# How diffusion tensor imaging findings in subconcussive head impacts correlate with head kinematic exposures: a systematic review and quantitative synthesis

**DOI:** 10.64898/2026.09.28.26364205

**Authors:** Maryam Tayebi, Christian John A. Saludar, William Schierding, Eryn Kwon, Joshua McGeown, Alan Wang, Justin Fernandez, Samantha Holdsworth, Vickie Shim

## Abstract

Collision and contact-sport athletes are repeatedly exposed to subconcussive head impacts or subclinical head acceleration events (HAEs), which are impacts that do not result in a clinical diagnosis of mild traumatic brain injury but may induce subtle neurological alterations. These effects may include structural, functional, and metabolic changes within the brain. Various exposure quantification techniques have been developed to characterise HAE exposure, while diffusion MRI enables assessment of brain microstructural organisation, with changes in diffusion-derived metrics associated with pathophysiological processes.

This systematic review was conducted in accordance with PRISMA guidelines to evaluate the use of various HAE exposure quantification techniques, exposure weighting approaches, and diffusion MRI in characterising the relationship between HAE exposure and brain microstructure in athletes. Across studies, longitudinal findings for fractional anisotropy (FA), mean (MD), axial (AD), and radial diffusivity (RD) were inconsistent, and substantial heterogeneity in exposure quantification methods and analyses was observed. However, quantitative analysis demonstrated a significant negative association between total subclinical HAE count across the season and FA, and a positive association with MD, suggesting cumulative microstructural alteration.

Exposure metrics—including risk-weighted, time-weighted, and finite-element-derived strain measures—showed stronger associations with diffusion MRI alterations than simple impact counts or peak acceleration measures. However, HAE exposure risk models are sport-specific and require broader validation for applicability across sporting cohorts. Future studies should reduce methodological heterogeneity, control for confounding variables, and refine exposure quantification and weighting approaches to better understand the relationship between repetitive subclinical HAE exposure and WM microstructural integrity.

## INTRODUCTION

Participation in sport is associated with benefits to physical, psychological, and social health [1,2]. However, in collision sports where purposeful collisions are a part of gameplay, injuries are common [3]. Rates of mild traumatic brain injury (mTBI) are higher in collision sports than in non-contact sports[4]. In recent years, interest in the risks associated with repetitive exposure to subconcussive impacts or head acceleration events (HAE) has increased. HAEs are events that cause head acceleration due to an external, short-duration collision or impact applied directly or indirectly to the head [5]. Hence, this includes head impacts and subconcussive impacts. HAE induces varying strain responses on neuronal tissue [2]. Repetitive exposure to HAE, even without mTBI diagnosis, has been associated with short-term or long-term negative effects on cognitive and physiological outcomes [5]. While most HAEs are ‘subclinical’, where force transmitted to the brain does not result in acute signs and symptoms consistent with diagnostic criteria for mTBI, these subclinical HAE may still induce neurophysiological changes [6]. For the remainder of this paper, we refer to HAE as those subclinical HAE that do not result to clinical diagnosis of mTBI, including subconcussive impacts. Studies have linked subclinical HAE in contact sport athletes to structural, functional, and metabolic alterations [7–10]. Since these events often cause no observable acute symptoms, subtle microstructural changes may go undetected, preventing proper recovery and increasing susceptibility to mTBI [11,12]. Recently, a study employed a position-exposure matrix to estimate lifetime HAE exposure, finding that duration of play and exposure-derived measures were significantly associated with post-mortem neuropathological changes [13]. This supports growing evidence linking exposure to subclinical HAE with long-term neurological impairments and higher rates of neurodegenerative diseases [14,15].

Traditional clinical neuroimaging methods (e.g. T1- or T2-weighted images) often fail to study the effects of subclinical HAE due to their limited sensitivity to the subtle changes on brain microstructure [16,17]. In contrast, advanced MRI techniques have shown promise in identifying diffuse axonal injuries and underlying structural alterations in the brain [18]. Diffusion tensor imaging (DTI) is widely utilised in neuroimaging research to investigate white matter (WM) changes [19]. DTI quantifies water diffusion in the brain, particularly the anisotropic diffusion along WM tracts, which is influenced by biological barriers such as myelin sheaths [20–22]. By measuring water movement in multiple directions, DTI derives three perpendicular eigenvalues used to compute diffusion indices [23]. DTI metrics include fractional anisotropy (FA; diffusion asymmetry), mean diffusivity (MD; average diffusion across three axes), axial diffusivity (AD; diffusion along the primary axis), and radial diffusivity (RD; diffusion perpendicular to the primary axis) [24]. Alterations in these metrics are linked to pathology—reduced FA and increased MD suggest demyelination and axonal degeneration, lower AD indicates disrupted axonal orientation, and higher RD is associated with myelin loss, or cytotoxic edema [25–28]. A brief overview of DTI metrics and their associated pathophysiology is presented in Figure 1. However, different WM changes can produce similar effects on diffusion parameters, emphasising the need to examine multiple metrics and their interrelationships [29,30].

**Figure 1.**
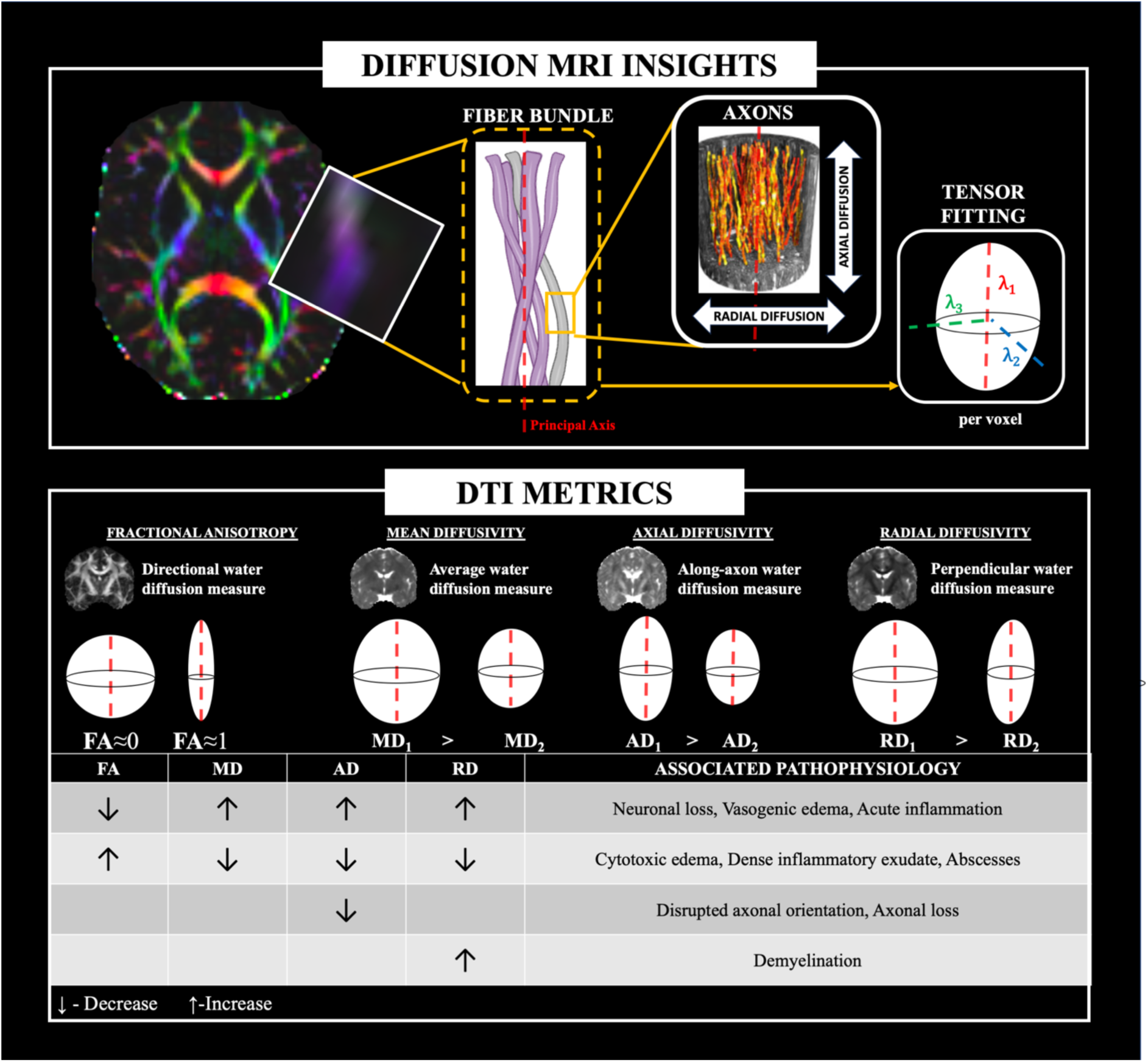
Brief overview of DTI metric derivation from diffusion MRI and the pathophysiology associated with observed directional changes. 3D visualisation of axons adapted from Figure 7A in Andersson et al. (2022), with permission from Elsevier.[37]

Reviews by Schneider et al., Tayebi et al., and Koerte et al. highlight significant inconsistencies in the directionality and regional distribution of these DTI changes as an effect of repetitive subclinical HAE exposure[31–33]. These inconsistencies are largely driven by heterogeneity in study design, sample cohort, and imaging protocol. In an attempt to address these limitations, Kwiatkowski et al. conducted a mega-analysis, integrating data from diffusion MRI studies on HAE while controlling for methodological heterogeneity [11]. Their findings revealed a lateralised pattern of diffusivity changes, with MD, AD, and RD increasing in central and brainstem regions but decreasing in the left hemisphere and frontal lobe. These results suggest strain-induced injuries in central and brainstem regions, and milder exertion-related effects in the frontal and left hemispheres. However, critically, there remains a lack of standardised measures of HAE exposure and a synthesis of the current methodological approaches to quantifying this exposure.

Recent research has emphasised the need for efficient methods to measure biomechanical forces from HAE kinematics to better study the subtle effects of HAE on brain microstructure [34]. To address this, in addition to counting HAE incidents or measuring linear and rotational acceleration, studies have adopted filtering techniques such as risk-weighted filtering[35] and time-weighted filtering [36]. Risk-weighted filtering evaluates cumulative exposure to repetitive HAE throughout a season, factoring in the risk associated with each event based on linear acceleration, rotational acceleration, or both. In contrast, time-weighted filtering considers both the recovery time and the brain’s ability to recover from repeated HAE by weighting impacts based on time between each impact or the time between the impact and the post-exposure assessment time. Tayebi et al., in their review, have also emphasised the necessity to incorporate kinematic measures in DTI studies to understand the relationship between the magnitude or number of HAE and changes in diffusion metrics.

Therefore, aim of this review is to collate all longitudinal studies that have employed DTI to assess the effects of subclinical HAE exposure on the brains of contact sport athletes. While most previous reviews have focused on the imaging aspect of these studies, this review has an added emphasis on the HAE quantification used and filtering techniques applied. Here, the quantitative analysis incorporates the most recent findings characterising subclinical HAE-induced injury by synthesising longitudinal DTI findings including the correlation of subclinical HAE exposure and DTI changes. In addition, it synthesises findings from longitudinal studies comparing contact-sport athletes exposed to subclinical HAEs with non-contact-sport players or mTBI-diagnosed subjects.

## METHODS

This systematic review included studies that utilised DTI to assess microstructural alterations in the brain resulting from repetitive exposure to subclinical HAE during participation in contact or collision sports. This literature followed the Preferred Reporting Items for Systematic Reviews and Meta-Analysis (PRISMA), consisting of the 27-item checklist and a four-phase flow diagram[38].

### Search strategy

Electronic databases, including PubMed, Scopus, ScienceDirect, and Web of Science, were searched to identify studies published up to December 2024 and written in English, based on title and abstract fields. In addition, reference lists of full-text papers, including systematic reviews related to the topic, were checked to identify additional eligible articles. Each database was searched using the specific keywords and Medical Subject Headings (MeSH) stated in Table 1. The utilisation of Boolean expressions guarantees that retrieved studies at least contain one combination of keywords in their title or abstract. The mTBI and concussion search term was used to identify papers that recruited subclinical HAE-exposed collision or contact sport athletes as controls. These subjects would have been exposed to subclinical HAE throughout the season or the study. In addition, ‘head impact’ was included to identify studies that used it rather than HAE. Papers retrieved from each database were uploaded to Rayyan QCRI [39], where paper screening was conducted.

**Table 1.** Keywords and search terms and inclusion and exclusion criteria.

|  |  |  |
| --- | --- | --- |
| Systematic search strategy |  |  |
| **Diffusion tensor imaging**<br>(diffusion tensor OR diffusion tensor imaging OR diffusion tensor magnetic resonance imaging OR diffusion tensor MRI OR diffusion tensor mris OR dti OR diffusion tractography OR tractography OR diffusion MRI) |  |  |
| **AND (mTBI OR concussion OR HAE OR subconcussive impact)**<br>(mild traumatic brain injury OR mtbi OR brain concussion OR brain concussions OR mild concussion OR mild concussions OR cerebral concussion OR sport concussion OR sport-related concussion OR src OR sports-related mild traumatic brain injury OR concussed athlete OR head acceleration events OR hae OR head impact OR rhi OR subconcussive OR subconcussive impact OR subconcussive injury OR post-concussive OR post-concussive injury) |  |  |
| **P.I.C.O. framework & study selection criteria** |  |  |
|  | **Inclusion criteria** | **Exclusion Criteria** |
| **Participations/population**<br>Active contact/collision sport male or female athletes with no clinical diagnosis of mTBI or concussion | Active contact/collision sport athletes with no clinical diagnosis of mTBI or concussion | Former, retired or inactive athletes<br>In vitro models of brain/neuronal/axonal injury<br>Animal models<br>Active athletes playing non-contact/collision sports |
| **Intervention/exposure**<br>Exposure to sported-related repetitive HAE or subconcussive impact | Sports-related collision like heading and scrum but does not result in mTBI/concussion. | Sports-related collision leading to diagnosis of mTBI/concussion |
| **Comparisons/controls**<br>Within-subject longitudinal data, pre-injury baseline measurement, neurologically normal control group, athletes diagnosed with sports-related mTBI, normative data derive from open-source databases | Presence of comparison data | Absence of comparison data |
| **Outcomes**<br>DTI utilised to characterise microstructural alteration due to repetitive subclinical HAE | Longitudinal studies with or without cross-sectional comparison and DTI was used to characterise microstructural alteration after repetitive exposure to HAE or subconcussive impact | DTI was not used |

### Inclusion criteria

All papers were reviewed based on their title and abstract to meet the following inclusion criteria: 1) longitudinal studies on humans; 2) subjects are active contact or collision sport players; 3) DTI or diffusion kurtosis imaging(DKI) was used to assess brain microstructural alterations due to subclinical HAE exposure. Papers with no full-text access even after accessing via the university library, records not written in English, and studies including retired athletes or former athletes were excluded from the analysis. The flow diagram of the database search and screening is shown in Figure 2.

**Figure 2.**
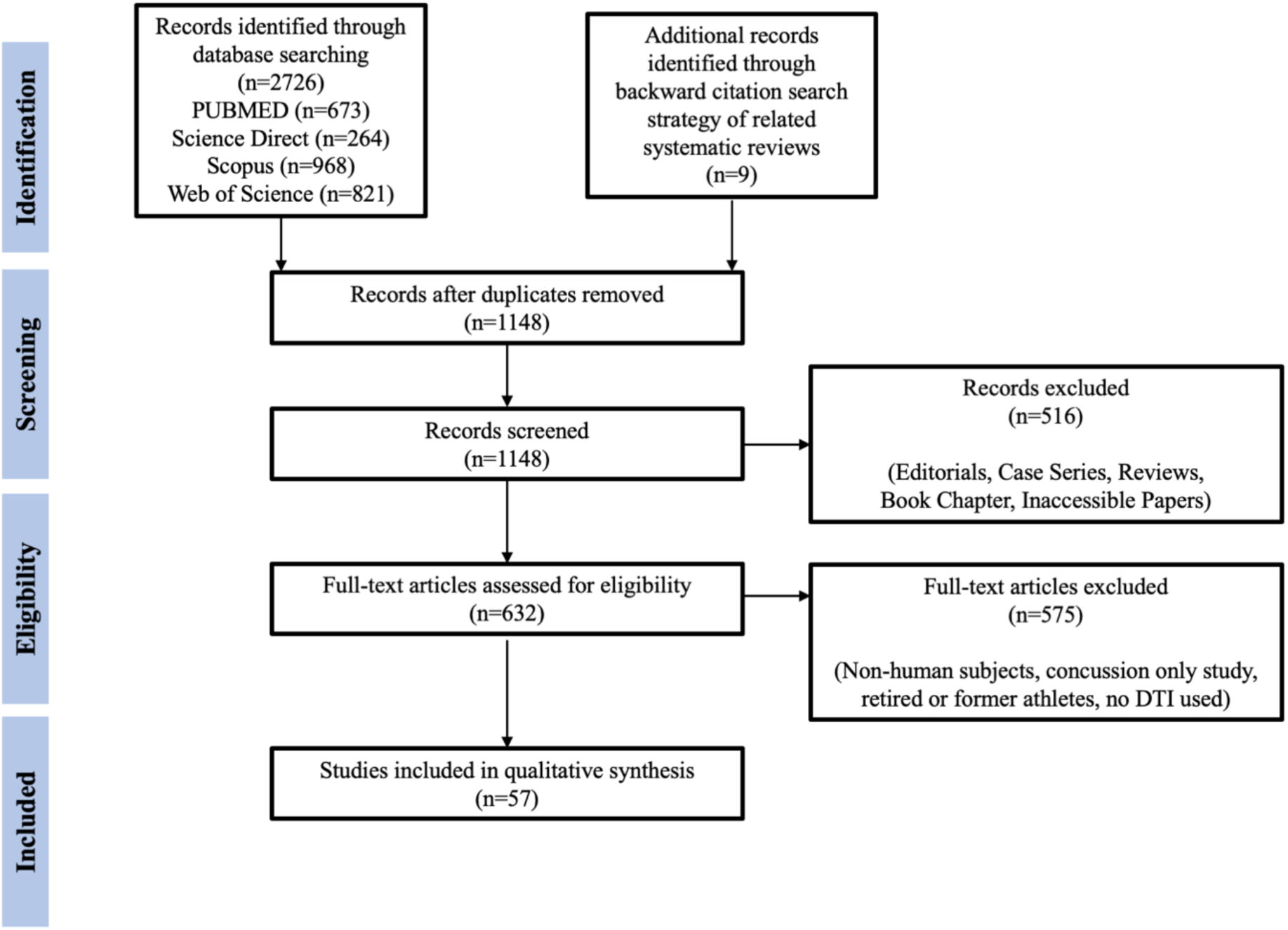
Flowchart for paper identification and selection.

In addition to the selection criteria for the database search, further criteria were imposed for inclusion into the quantitative analysis: (1) studies measured DTI change before(pre-season) and after (post-season) time points, (2) studies explicitly reported sufficient statistical data on bivariate correlation of DTI change with subclinical HAE exposure including its direction, (3) DTI change was reported as a longitudinal change and not as a cross-sectional comparison of control and experimental group, and (4) studies with rating based on risk of bias assessment greater than Quartile 1 (83%).

### Risk of bias assessment

A risk-of-bias assessment was conducted to identify flaws in the study methodology. The modified National Institute of Health (NIH) Quality Assessment Tool for Observational and Cohort and Cross-Sectional Articles utilised by Lees et al. was employed [40]. This assessment tool was further modified to fit the current aim of the review. Details of the modifications are available in the attached copy of the questionnaire in the supplementary materials - A. Key modifications included 1) sufficient reporting of subject demographics, including recruitment and inclusion or exclusion criteria; 2) in-depth quality assurance of reporting DTI findings, including direction of change and insignificant results; and 3) making the assessment tool applicable to assess the quality of various DTI study designs particularly accounting for longitudinal studies with and without cross-sectional comparison performed and studies with longitudinal cohort, yet only performed cross-sectional comparison with controls . Similar to Lees et al. (2021), studies were grouped into three quality categories based on weighted ratings and quartile (low [quartile 1], medium [quartile 2], and high [quartile 3-4]).

The modified assessment tool included 12 questions related to study aims and objectives, sample size, study design, validity of utilised measures of exposure, quality of DTI acquisition and analysis, sufficiency of reporting study findings, and appropriate consideration of confounding factors. Specifically, question 10 aimed to assess the DTI protocol and analysis in detail.

### Data extraction

The following information was extracted from the papers included in the review: title, author, year of publication, subject characteristics (population size, age group, sex, and sports played), DTI parameters, DTI data analysis methods, DTI metrics used for analysis, HAE exposure tools and measures (if applicable), and clinical and psychological tests conducted. In instances where the included study cited previous work as a reference for its methods, the cited reference was accessed, and available data were extracted. No interpolation of missing data was performed, and only reported data or quantitative values included were extracted.

Study findings of interest included pre-season to post-season changes in DTI metrics (FA, MD, AD, and RD), as well as the association between cumulative HAE and DTI changes. For studies included in the quantitative synthesis, effect sizes were extracted or computed for all available data, including reported correlation coefficients between longitudinal DTI changes and kinematic exposure metrics. Consistent with Lees et al.[40], effect sizes were derived from group means and standard deviations, t-values, or p-values, depending on the data reported.

For studies reporting changes in DTI metrics or correlation coefficients across multiple regions of interest (ROIs), a single study-level effect size was obtained by pooling regional effects. Cohen’s d was used to quantify effect sizes (standardised mean differences) for longitudinal changes in DTI metrics[41]. Effect sizes for associations were based on pooled bivariate correlation coefficients across studies [42]. Due to the limited number of studies examining associations between HAE measures and DTI outcomes, only correlations between cumulative impact count (i.e., the total number of HAE exposures across a season) and FA or MD were included in the quantitative synthesis. Effect sizes were interpreted as small (0.2), medium (0.5), and large (0.8).

Findings of interventional studies were excluded from analysis unless separate statistics were reported for the non-intervention control group. Results from a study reporting no directionality of increase or decrease for DTI changes or positive or negative correlations were excluded. In addition, for multiple studies using the same dataset and reporting statistics for the same metric, only the most recent paper publication was included to prevent unit-of-analysis problems in quantitative synthesis. In contrast, separate effect size estimates were derived for each contact sport group with independently reported statistics within a single study. Quantitative synthesis was conducted using the R(“R version 4.4.1”) statistical software [43] and the ‘meta’ package [44].

## RESULTS

### Search results

The literature search through the electronic databases yielded 2726 records. An additional nine papers were found through a backward citation search strategy. After the removal of duplicates, 1148 papers were left. From these, 516 papers were excluded as they were not original articles, were case studies, were editorials, were review papers, or were irrelevant to the topic of the present review. The eligibility of the remaining 632 papers was assessed by considering the inclusion and exclusion criteria. A total of 57 studies were included in the final qualitative analysis (Figure 2). Collected data from each study are summarised in supplementary material - B

### Study characteristics

#### Level of play and sports

An overview of study characteristics is presented in Figure 3, including stratification by level of play, sport, and sex.

**Figure 3.**
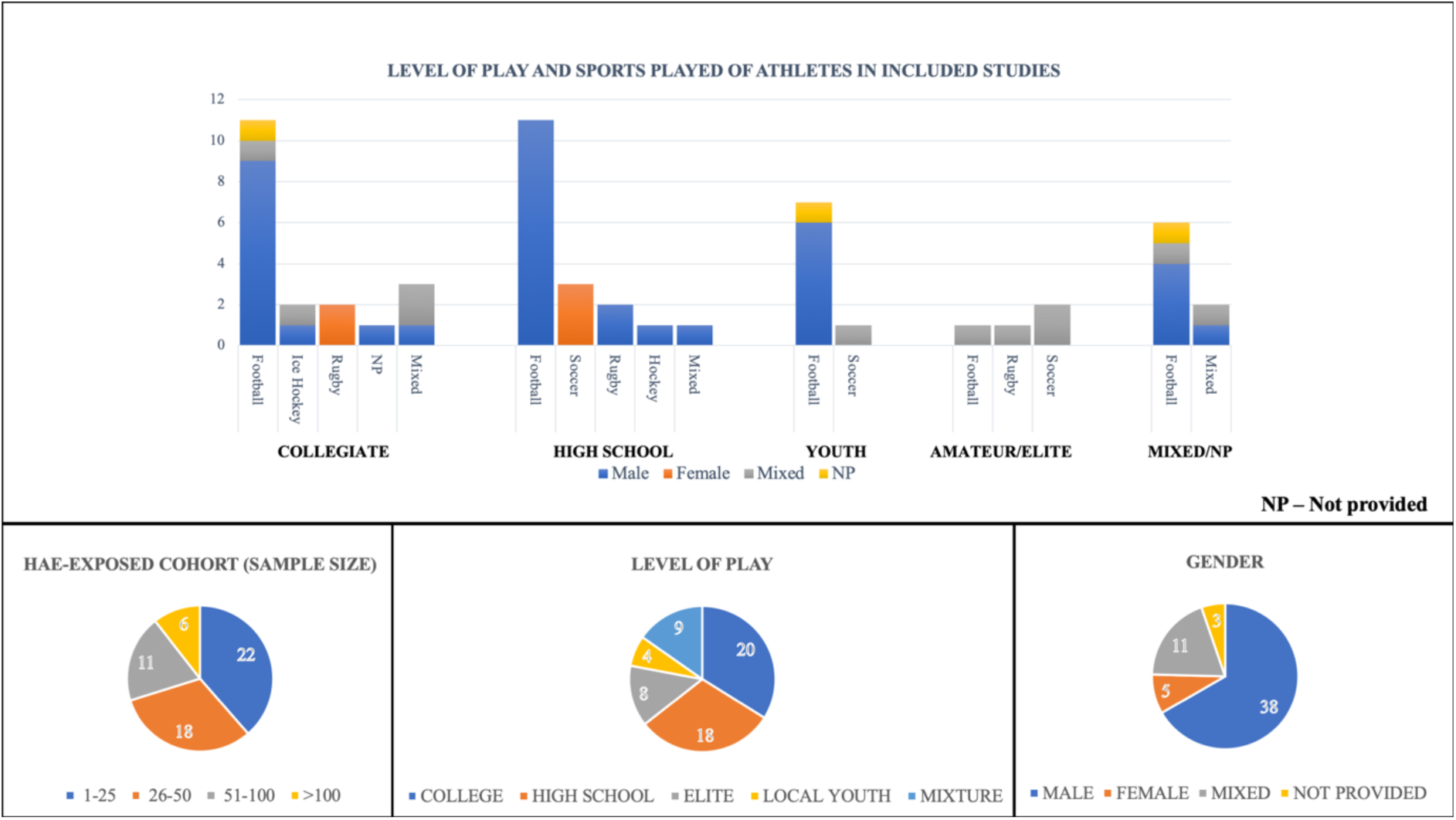
Subject demographics in included studies stratified according to level of play, sports, sample size, and sex.

Collegiate athletes were the most frequently studied population (n = 20), followed by high school athletes (n = 18), amateur or elite athletes (n = 8), and youth athletes competing at the local level (n = 4). Nine studies included mixed athletic age groups. One study specifically compared youth and high school athletes to examine differences in brain microstructural alterations associated with subclinical HAE exposure [45]. Regarding sport representation, the majority of studies (n = 36) involved American football athletes. Other sports included mixed-sport cohorts (n = 6), soccer (n = 6), rugby (n = 5), ice hockey (n = 2)[46,47], and field hockey (n = 1)[48]. Two studies did not specify the sport of the included athletes[19,49].

#### Sample size and sex

In terms of sample size, most of the studies. (N=22) had a sample size of repetitive subclinical HAE-exposed athletes of less than 25. Only six studies had a sample size exceeding 100. The majority of included studies evaluated male athletes only (N=38). A fraction of included studies included mixed-sex subjects (N=11), while five studies focused on female athletes only. Three did not specify the gender of the included subjects[8,36,50].

#### Study design and time point of measure

Most studies (N=43) scanned athletes at the beginning (pre-season) and conclusion (post-season) of their competitive seasons. Within these, eight studies added scans in the pre-season and post-season to include the off-season (N=4)[8,51–53], mid-season (N=4)[48,54–56], or before training camp (N=1)[57]. On the other hand, nine studies scanned subclinical HAE-exposed subjects for mTBI symptom monitoring (acute, sub-acute, or chronic) in mTBI-diagnosed subjects, while three studies performed imaging at baseline and at least 1 year after. One study each did a scan before and after a weekend game [58] or a single game[59]. Of the studies focusing on longitudinal changes in subclinical HAE-exposed subjects, nine further grouped subjects into collar or non-collar groups (N=5)[48,53,60–62], utilised sports helmets (N=1)[63], sex (N=1)[47], impact exposure (N=1)[57], or strain severity (N=1)[64].

Twenty-six studied compared subclinical HAE-exposed athletes with control and experimental groups (i.e. mTBI-diagnosed subjects). Summary results of studies testing the difference between non-contact sport controls and contact sport athletes, or contact sport athletes and mTBI-diagnosed athletes, are presented in Supplementary C. Although caution should be exercised in interpreting these graphs, as they do not represent the spatial extent of significant differences and may be based on a single significantly different region among all regions studied.

#### Imaging acquisition and analysis

Diffusion MRI was acquired in all studies using a 3.0 Tesla scanner. Among studies reporting acquisition parameters, 37 used a single b-value, while 18 used multiple b-values. FSL was the most used pre-processing package across all included studies.

Most studies (N=43) employed voxel-based analysis, with 24 implementing Tract-based Spatial Statistics. Twenty-six studies applied ROI-based analysis, as a single approach or in addition to a voxel-based approach. One conducted structural connectivity analysis[53], while one utilised Fixel-based analysis[65].

A novel subject-specific analysis method was proposed by Asselin et al., referred to as the SPREAD algorithm, which improves the statistical power of group-based analysis by focusing on varying subject-specific baseline DTI parameters and disease progression [66]. In the field of ROI analysis, Tayebi et al. proposed tract morphing, which generates subject-specific 3D models matching subject-specific tracts or any region of interest, which improves comparability and discernibility of DTI metrics in differentiating cases and controls [56].

Two studies used machine learning to classify subclinical HAE-exposed subjects based on impact exposure[50] or subclinical HAE-exposed athletes from non-collision sport controls[67]. These highlight the applicability of machine learning applied to DTI in better understanding the microstructural alterations caused by repetitive subclinical HAE exposure.

### Diffusion tensor imaging

Of the 43 studies that performed pre-season-to-post-season scanning, 35 explicitly compared DTI metrics within the season. The remaining eight either focused on machine learning classification of degree of exposure (N=2)[50,67], only performed cross-sectional comparison with controls (N=2)[19,56], pre-season to mid-season or other timepoint comparison (N=1)[48], structural connectivity graph theory measures (N=1)[53], DKI metrics (N=1)[68], or analysis between two successive seasons and not within a season (N=1)[69].

Among the 35 studies focused on pre-post comparisons, FA was studied in all, MD in 26, and both AD and RD in 17. A summary of reported changes from pre-season to post-season is presented in Table 2. An observable, inconsistent trend across DTI metrics in these studies is observable.

**Table 2.** Summary table on directional change observed in included studies from pre-season to post-season.

| NUMBER | STUDY | ANALYSIS METHOD | HAE EXPOSURE | FA | MD | AD | RD |
| --- | --- | --- | --- | --- | --- | --- | --- |
| 1 | Asselin et al. (2020)[66] | SPREAD | HITS (<10 g) | ↑↓ (Mostly decrease) | -- | -- | -- |
| 2 | Bahrami et al. (2016)[70] | ROI-Based | HITS (Focused on RWE-CP) | ↑↓ (Mixed) | -- | -- | -- |
| 3 | Barber Foss et al. (2019)[45] (HS Group) | Voxel-based ROI for localisation | GForceTracker | NS | ↓ | ↓ | ↓ |
|  | Barber Foss et al. (2019) (Youth) | Voxel-based ROI for localisation | GForceTracker | ↑ (If concussed athlete is excluded) | NS | ↓ | NS |
| 4 | Bazarian et al. (2012)[71] | Voxel-based with ROI for localisation and ROI-based | Self-reported Hits | ↑↓ (*) | ↑↓ (*) | -- | -- |
| 5 | Bazarian et al. (2014)[51] | Voxel-based with ROI for localisation | HITS (>10 g) | ↓ (*) | ↑↓ (*) | -- | -- |
| 6 | Champagne et al. (2019)[57] | Voxel-based on a ROI | GForceTracker (>15g) | ↓ | -- | -- | -- |
| 7 | Chun et al. (2022)[54] (Team 1) | ROI-based (~40 tested) | HITS (thr not stated) | ↑ (2 ROIS) | -- | -- | -- |
|  | Chun et al. (2022) (Team 2) | ROI-based (~40 tested) | HITS (thr not stated) | ↓ (1 ROIS) | -- | -- | -- |
| 8 | Davenport et al. (2014)[72] | Voxel-based | HITS (>10 g) | Not reported (*) | Not reported (*) | -- | -- |
| 9 | DiCesare et al. (2020)[73] | Voxel-based | XPatch (>10g) | Not reported | Not reported | No reported | Not reported |
| 10 | Diekfuss et al. (2021)[62] | Voxel-based ROI for localisation | CSx Sensor (>20 g) | ↑ (Non-collar) | ↓ (Non-collar) | ↑↓ (Non-collar) | ↓ (Non-collar) |
| 11 | Diekfuss et al. (2021)[63] | Voxel-based ROI for localisation | CSx Sensor (> 10 g) | ↑ (HRank Helmet) | ↓ (LRank Helmet) | ↑↓ (LRank Helmet)<br>↑ (HRank Helmet) | ↓ (LRank Helmet) |
| 12 | Gong et al. (2018)[74] | Cluster-wise ROI-based | HITS (>10 g) | NS | ↑ (****) | -- | -- |
| 13 | Goubran et al. | Tract Profiling | HITsp | ** | ** | No reported | ** |
|  | (2023)[75] |  |  |  |  |  |  |
| 14 | Holcomb et al. (2021)[64] | Voxel-based ROI for localisation | HITS (thr not stated) | Not reported | Not reported | -- | -- |
| 15 | Jang et al. (2019)[76] | Voxel-based and ROI-based | HITS & xPatch (>20 g) | ↑ (Mean of altered mask) | NS | -- | -- |
| 16 | Koerte et al. (2012)[77] | Voxel-based | None | NS | -- | ↑ | ↑ |
| 17 | Kuzminski et al. (2018)[78] | Voxel-based and ROI-based | HITS (>10 g) | NS | -- | -- | -- |
| 18 | Lao et al. (2015)[49] | Vertex-wise | None | NS | NS | -- | -- |
| 19 | Manning et al. (2020)[79] | ROI-based | Head Sensor (GFT3) | ↓ (Brainstem) | ↑ (Brainstem) | ↑ (Brainstem) | ↑ (Brainstem) |
| 20 | Marchi et al. (2013)[8] | Voxel-based | Head Hit Index (HHI) | Not reported | Not reported | -- | -- |
| 21 | Mayinger et al. (2018)[52] | Voxel-based | None | ↑ | -- | NS | NS |
| 22 | McAllister et al. (2013)[80] | ROI-based | HITS (>14.4 g) | *** | *** | -- | -- |
| 23 | Merchant-Borna et al. (2016)[81] | Voxel-based | HITS (>10 g) | ↑ ↓ (*) | -- | -- | -- |
| 24 | Merz et al. (2020)[82] | ROI-based | HITS (>14.4 g) | NS | NS | -- | -- |
| 25 | Miller et al. (2022)[83] | Voxel-based | HITS (>10 g) | Not reported | Not reported | -- | -- |
| 26 | Mund et al. (2024)[84] | Voxel-based and ROI-based | Heading | NS | NS | NS | NS |
| 27 | Myer et al. (2016)[60] | Voxel-based ROI for localisation | GForce Tracker (>20 g) | NS | ↓ (Non-collar) | ↓ (Non-collar) | ↓ (Non-collar) |
| 28 | Myer et al. (2019)[61] | Voxel-based ROI for localisation | xPatch | NS | ↓ (Non-collar) | ↓ (Non-collar) | ↓ (Non-collar) |
| 29 | Nilsson et al. (2019)[85] | ROI-based | Shockbox (>30 g) | NS | -- | -- | -- |
| 30 | Puvvada et al. (2021)[36] | Voxel-based | HITS (thr not stated) | Not reported | Not reported | -- | -- |
| 31 | Schranz et al. (2018)[86] | ROI-based | None | ↑ (MRS Voxel) | NS | NS | ↓ (MRS Voxel) |
| 32 | Slobounov et al. (2017)[10] | **Voxel-based** and ROI-based | BodiTraK (>25g) | NS | NS | NS | NS |
| 33 | Sollmann et al. (2018)[47] | Voxel-based ROI for localisation | None | ↓ (Female) | ↑ (Female) | ↑ (Female) | ↑ (Female) |
| 34 | Tayebi et al. (2024)[55] | Voxel-based ROI for localisation | HITIQ (>59g) | NS | NS | NS | NS |
| 35 | Yuan et al. (2018)[87] | Voxel-based ROI for localisation | GForce Tracker (>10 g) | ↑ (Non-collar) | ↓ (Non-collar) | ↓ (Non-collar) | ↓ (Non-collar) |

\* Counted % of significantly changed voxels based on permutation testing, bootstrapping, or z-score based on controls \*\* Attenuated longitudinal trajectory compared with non-contact sport controls \*\*\* Main effect of group x time, but no explicit statement of significant change \*\*\*\* Grey Matter regions NS Not Significant -- Did not include in the analysis

The directional change reported in this paragraph is either driven by an ROI analysis or a voxel cluster and may not reflect widespread change. For FA, 13 studies reported no significant longitudinal change, 8 reported an increase, 5 reported a decrease, and 3 reported bidirectional change. MD, on the other hand, was observed to reveal no significant change in 8 studies, a significant decrease in 6, an increase in 3, and a bidirectional change in 2. Furthermore, AD revealed no significant change in 5 studies, decreases in 5 studies, increases in 3, and bidirectional changes in 2. Lastly, RD showed significant decreases in 7 studies, significant increases in 3, and no significant change in 4. The reported bidirectional changes are partly due to the method used, which quantifies the percentage of WM voxels with significant increases or decreases, either via bootstrapping, the SPREAD algorithm, or z-scored comparisons with the normative control’s mean and standard deviation. Moreover, a few of the studies did not explicitly report the direction of change, but reported only the correlation of DTI metric changes with HAE kinematics.

Region exhibiting pre-post season changes are illustrated in Figure 4. The most reported region with season longitudinal alteration in DTI metric/s is in the right superior longitudinal fasciculus (N=10) and corpus callosum (Body=8, Splenium=7, Genu=6).

**Figure 4.**
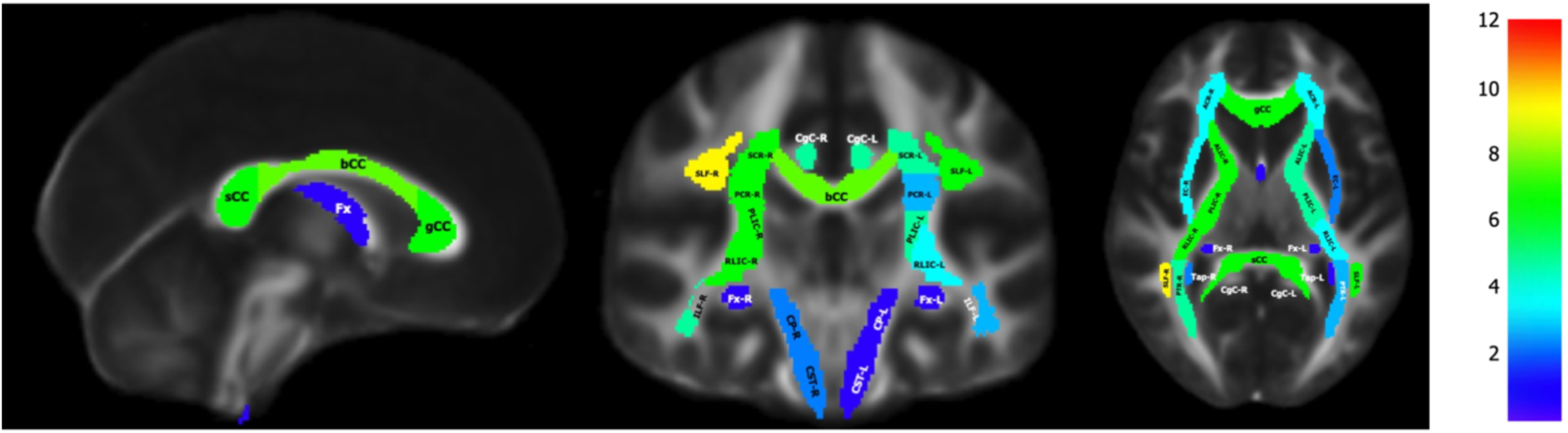
Colour coding on ICBM-DTI-81 white-matter labels brain atlas showing the number of papers reported significant longitudinal pre-season to post-season change in athletes exposed to repetitive subclinical HAE. Color bar represents “number of papers”. bCC (Body of Corpus Callosum), gCC (Genu of Corpus Callosum), sCC (Splenium of Corpus Callosum), CP (Cerebral Peduncle), CG (Cingulum), ACR (Anterior Corona Radiata), PCR (Posterior Corona Radiata), SCR (Superior Corona Radiata), CST (Corticospinal Tract), EC (External Capsule), Fma (Forceps Major), Fmi (Forceps Minor), IFOF (Inferior Fronto-Occipital Fasciculus), ILF (Inferior Longitudinal Fasciculus), ALIC (Anterior Limb of Internal Capsule), PLIC (Posterior Limb of Internal Capsule), RLIC (Retrolenticular Limb of Internal Capsule), SFOF (Superior Fronto-Occipital Fasciculus), SLF (Superior Longitudinal Fasciculus), Tap (Tapetum), ATR (Anterior Thalamic Radiation), PTR (Posterior Thalamic Radiation), STR (Superior Thalamic Radiation), UF (Uncincate Fasciculus)

Of the studies that compared the subclinical HAE-exposed group with other groups cross-sectionally, 20 studies compared subjects subclinical HAE-exposed with normal controls (non-contact sport athletes, orthopaedic injured, or non-athletic groups), 14 compared with mTBI-diagnosed subjects, and 5 included all three groups: mTBI-diagnosed, subclinical HAE-exposed, and non-collision athlete or neurotypical controls.

In studies comparing subclinical HAE-exposed subjects with normal controls, FA was studied in 20, MD in 17, AD in 8, and RD in 8. In studies comparing them with mTBI-diagnosed subjects, FA was studied in 14, MD in 14, AD in 10, and RD in 10.

### HAE exposure quantification

A summary description of all methods of quantifying HAE exposure is presented in Table 3. Generally, tools used to quantify HAE can be grouped into helmet-based sensors, patch-based/skin-mounted sensors, instrumented mouthguards, or questionnaire-based exposure estimation. Overall, HAE exposure was mostly quantified using cumulative impact count or the frequency of hits or headers in 27 studies.

**Table 3.** Various tools used in included studies to record HAE intensity.

| GROUPING | TOOL | DESCRIPTION |
| --- | --- | --- |
| Helmet-based Sensors | Head Impact Telemetry System (HITS) | Instrumented helmets fitted with a modified helmet liner that positions an array of 6 single-axis accelerometers against the head to enable in-vivo head acceleration measurements[80] |
|  | GForceTracker | A device affixed to the inside of each football helmet and records linear acceleration and rotational velocity by measuring three axes of linear acceleration and three axes of angular acceleration[48] |
|  | BodiTrak | A helmet-based sensor comprise of elastic fabric with pressure monitors and impact sensors that record both linear acceleration and location of impact[10] |
|  | ShockBox | A commercially available product that uses binary force switches that delineates impact to the front, rear, top, and lateral aspects of the helmet[85] |
| Patch-based/Skin-mounted Sensors | CSx Sensor | The CSx sensor is comprised of a tri-axial accelerometer and gyroscope that quantifies linear and rotational acceleration. It is affixed to the left mastoid process of the subject[62] |
|  | XPatch | A small, durable skin-worn sensor affixed to the mastoid process behind the athlete's ear via adhesive. The device records tri-axial linear acceleration and angular velocity to track head kinematics resulting from impacts.[61,73] |
| Instrumented Mouthguards | HitIQ® Nexus A9 | A bespoke instrumented mouthguard that is molded to fit the oral cavity of each player and records linear and angular acceleration[55] |
| Questionnaire-based Exposure Estimation | High Position-based Impact Risk (HITsp) | It is a composite measure that quantifies a player's risk based on hit impact rotational and linear acceleration, quantity, duration, and location for other players of the same position[75] |
|  | Head Hit Index (HHI) | A tool to quantify occurrence of head hits. This was used to track the frequency and severity of significant collision a player experienced during each game[8] |
|  | HeadCount | Structured, web-based questionnaire that estimates the heading over the prior time period based on a structured questionnaire that assess exposure during practice and games[88] |
|  | Years of Contact Sport Exposure | Quantifies the number of years of participating in contact sport of a subject[89] |
|  | Self-reported Hits | Record number of hits |

Thirty-six studies utilised kinematic sensors and questionnaires to quantify subjects’ exposure to HAE. Eighteen studies utilised the Head Impact Telemetry System (HITS), five used the GForceTracker, three used the CSxPatch, two used the HitIQ instrumented mouthguard, and the rest used XPatch, BodiTrak, ShockBox, and questionnaires. Various HAE magnitude thresholds were reportedly used by studies with most setting it at 10g (N=10) while other varied at 14.4g[80,82], 20g[62,76], and 59g[55].

Using the aforementioned tools, studies derived various metrics, including device measures (mean, peak, or cumulative linear or angular acceleration/velocity) for 23 studies. Using these measures, studies utilised developed weighting techniques, including risk-weighted exposure (RWE) used in 7 studies and time-weighted exposure including time between hits (TBH) or time until assessment (TUA) in 3 studies[36,66,81]. A description of these filtering techniques are presented in Table 4. In addition, three studies have used strain-based measures from finite element (FE) analysis using amplified MRI[57] or simulating recorded impacts on the Atlas-based Head Model[64,83].

**Table 4.**
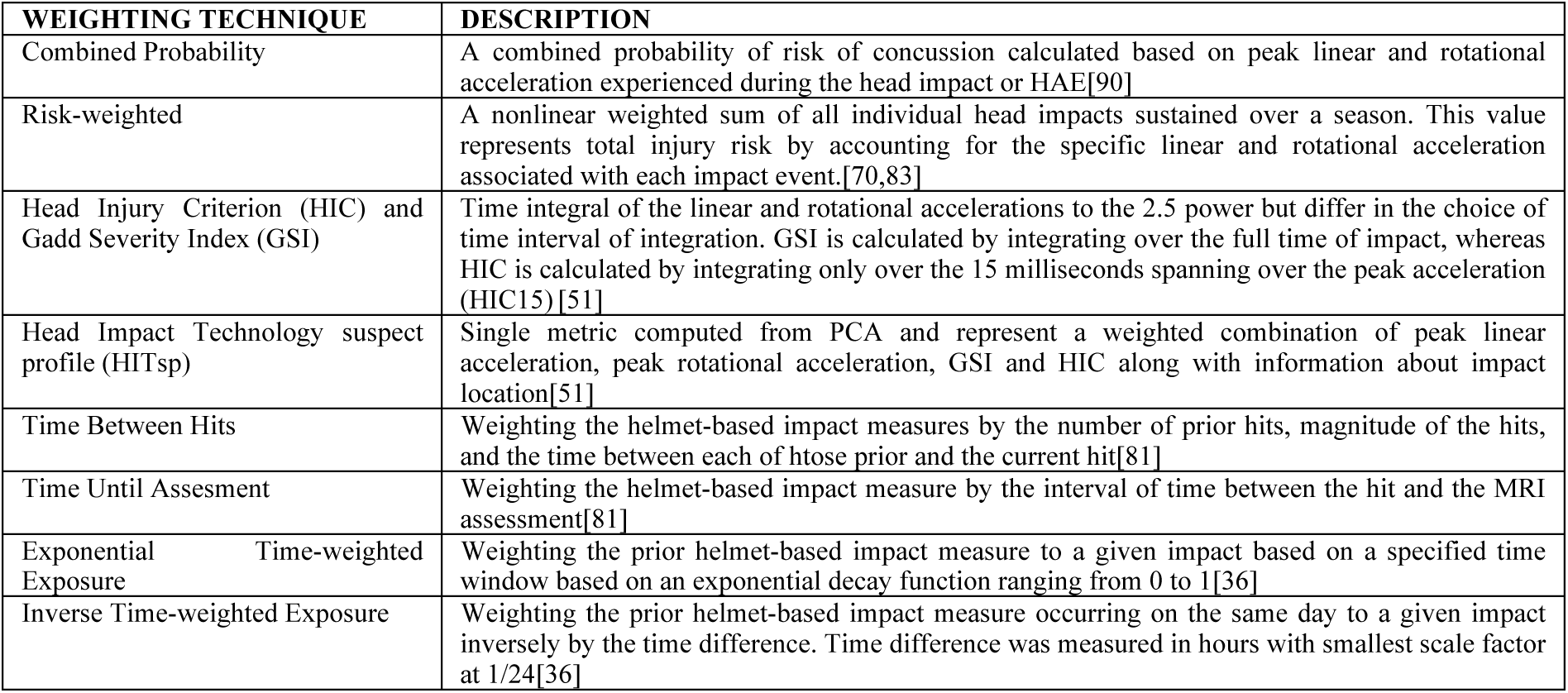
Varying weighting applied to head acceleration events in included studies.

Among studies that compare pre-season to post-season, 21 found a correlation between DTI metric changes and HAE exposure metrics. This comprises 11 studies that find seasonal changes in DTI metrics with cumulative impact count, and 14 that use device measures, including weighted or unweighted linear acceleration, rotational acceleration, or combined probability. Furthermore, 4 studies found a correlation with head injury criteria (HIC), Gadd severity index (GSI), and head impact technology suspect profile (HITsp) with DTI changes[51,66,80,81], and 2 studies found the same, but with FE stress and strain metrics[64,83]. However, these correlations is with high heterogeneity, as some studies reported only a correlation between HAE exposure and specific ROI/s[45,55,66,70–72,78,80,85,91] or widespread regions based on voxel-based analysis[66,71,72,81,83,91]. These results are summarised in Table 5.

**Table 5.**
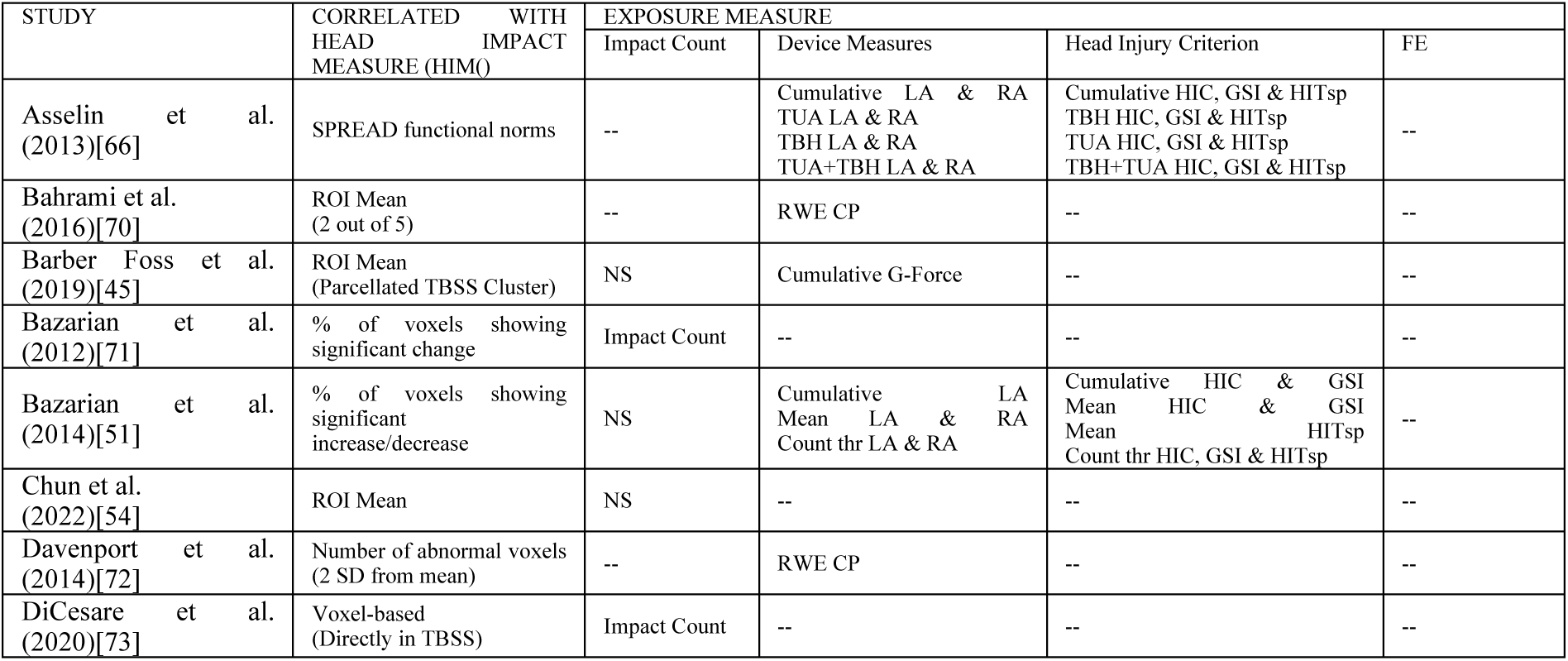
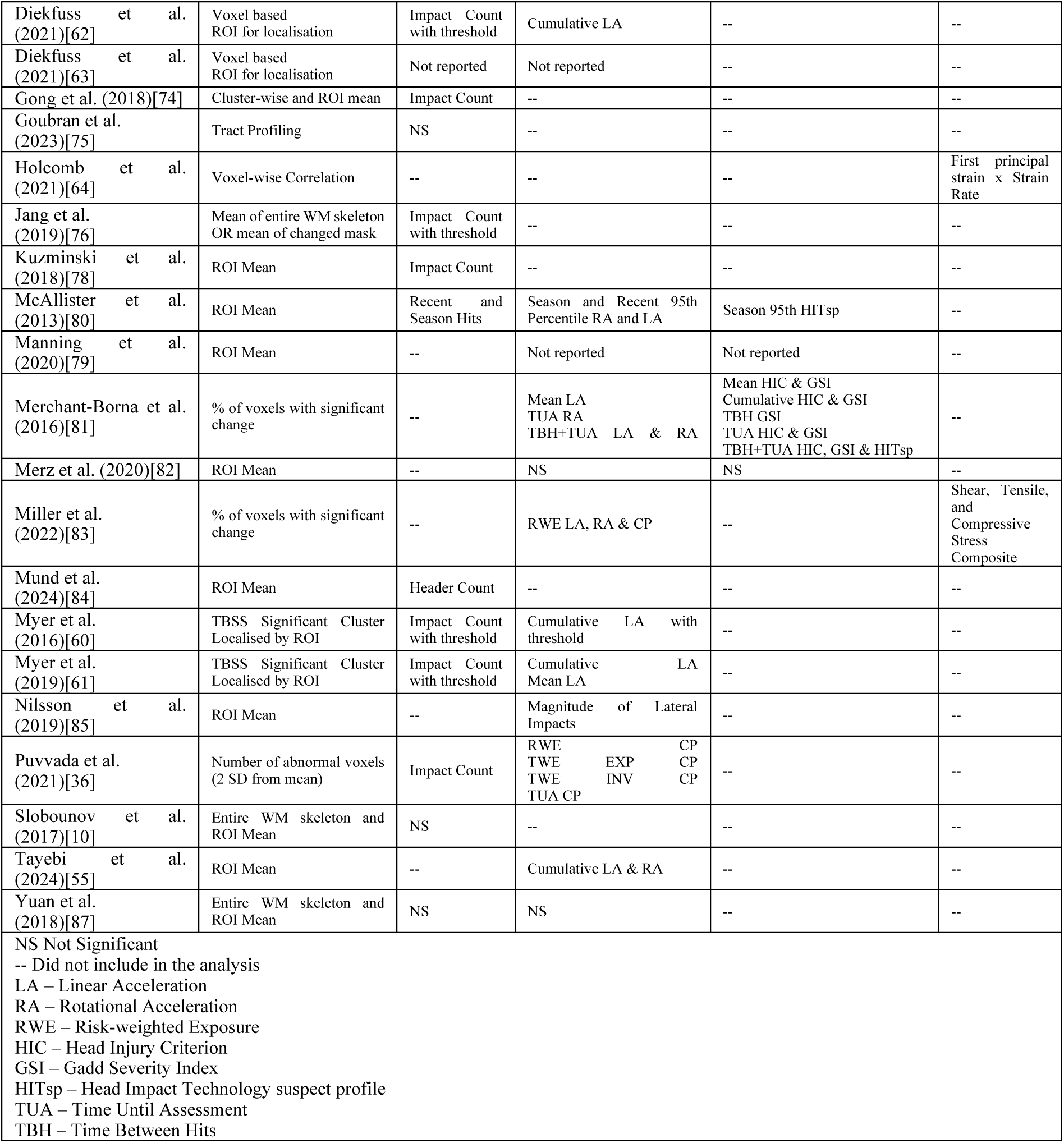
HAE metrics revealed to be correlated with pre-season to post-season DTI metric changes in included studies.

Among studies reporting a correlation between subclinical HAE exposure and pre-season-to-post-season DTI changes in ROIs, the right superior longitudinal fasciculus (N=6) and the corpus callosum (body=3, genu=4, splenium=5) were most often implicated. All other regions are illustrated in Figure 5.

**Figure 5.**
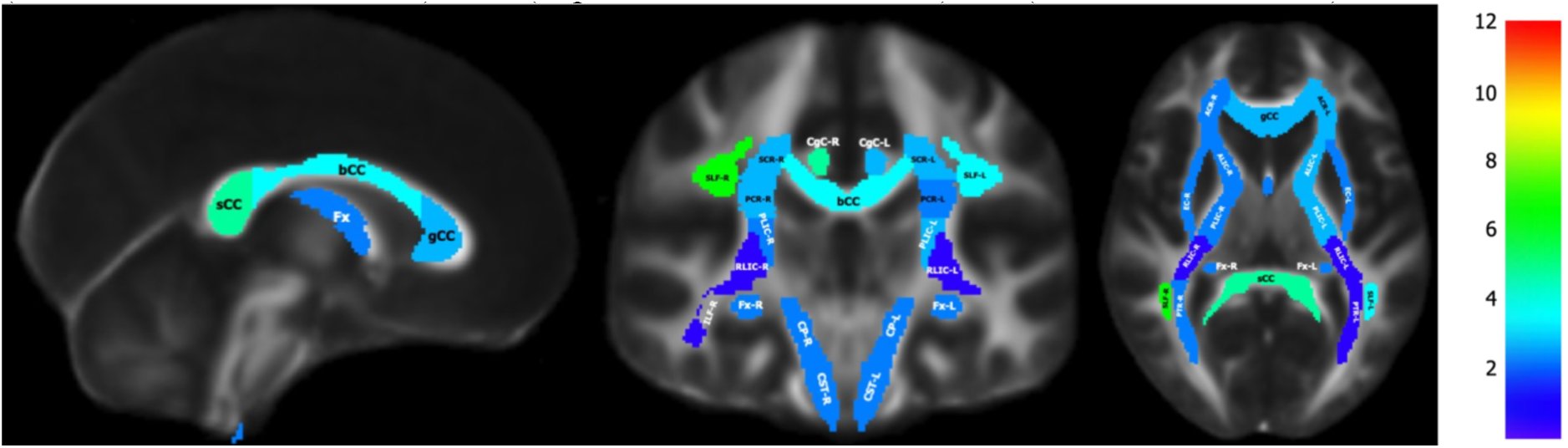
Colour coding on ICBM-DTI-81 white-matter labels brain atlas showing the number of papers reported significant longitudinal pre-season to post-season change correlation with HAE exposure in athletes exposed to repetitive HAE. Color bar represents “number of papers”. bCC (Body of Corpus Callosum), gCC (Genu of Corpus Callosum), sCC (Splenium of Corpus Callosum), CP (Cerebral Peduncle), CG (Cingulum), ACR (Anterior Corona Radiata), PCR (Posterior Corona Radiata), SCR (Superior Corona Radiata), CST (Corticospinal Tract), EC (External Capsule), Fma (Forceps Major), Fmi (Forceps Minor), IFOF (Inferior Fronto-Occipital Fasciculus), ILF (Inferior Longitudinal Fasciculus), ALIC (Anterior Limb of Internal Capsule), PLIC (Posterior Limb of Internal Capsule), RLIC (Retrolenticular Limb of Internal Capsule), SFOF (Superior Fronto-Occipital Fasciculus), SLF (Superior Longitudinal Fasciculus), Tap (Tapetum), ATR (Anterior Thalamic Radiation), PTR (Posterior Thalamic Radiation), STR (Superior Thalamic Radiation), UF (Uncincate Fasciculus)

### Quantitative synthesis

This section presents the quantitative synthesis performed in this study. Caution shall be exercised in its interpretation, as significant heterogeneity in reporting (e.g. widespread (WS) and ROI-based) was observed across studies.

#### Longitudinal changes in DTI metrics

Computed effect sizes (Cohen’s d) quantifying pre-season to post-season changes in DTI metrics are presented in Figure 6. Following the removal of overlapping cohorts, the remaining records provided data for observed change from pre-season to post-season in FA (n = 10), MD (n = 7), AD (n = 6), and RD (n = 7).

**Figure 6.**
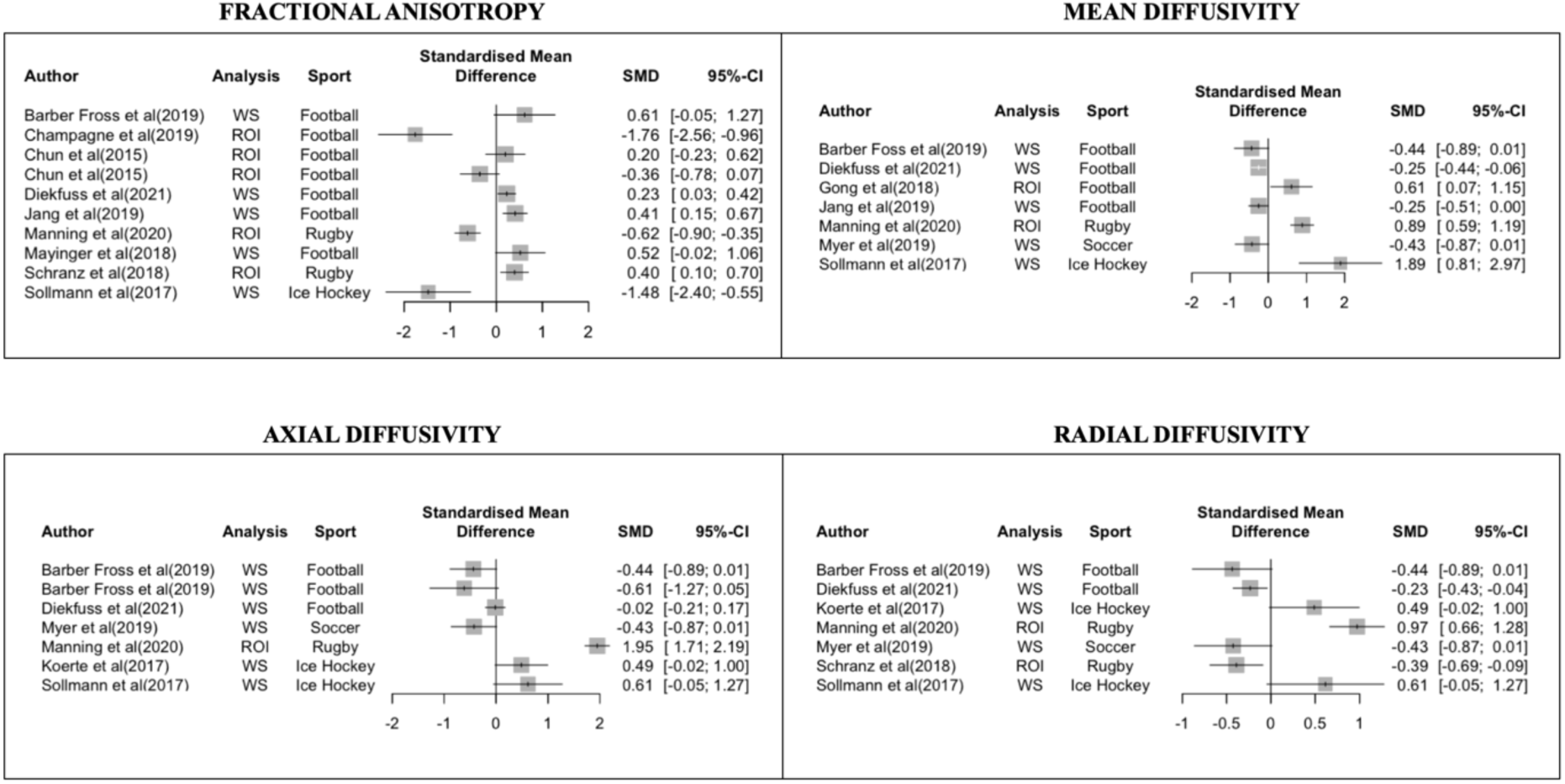
Forest plot of longitudinal pre-season to post-season DTI metric findings. Cohen’s d quantifying pre-season to post-season changes according to DTI metric. SMD (Standardised Mean Difference), 95%-CI (Confidence Interval)

FA, MD, AD, and RD demonstrated a high degree of variance. For FA, large effect sizes revealing a decrease were observed in two studies (d=-1.76,-1.48)[47,57], while moderate effect sizes were reported for both an increase in two studies (d=0.61,0.52)[45,52] and a decrease in one (d=-0.62)[79]. Remaining studies reported small increases and decreases. For MD, large increases were identified in two studies (d=1.89,0.89)[47,79], with a moderate increase observed in one study(d=0.61)[74]. Remaining studies yielded small decreases in MD, with effect sizes ranging from -0.25 to -0.44.

AD revealed a large increase in one study (d=1.95)[79], while moderate effect sizes were observed as decreases in one study (d=-0.61)[45] and as an increase in one (d=0.61)[47]. The remaining results were characterised by a small increase, with mostly showing decreases. Lastly, RD metric showed a large increase in one study (d=0.97)[79] . Moderate increase was observed in one study (d=0.61)[47], while majority of remaining studies demonstrated small decreases. It should be noted that these changes reveal heterogeneity not only in direction, but also the method used as split into widespread or ROI-based.

#### Impact count correlation with FA and MD

Given the very sparse studies reporting correlations between pre-season and post-season changes in DTI metrics and HAE exposure, the quantitative synthesis was confined to the correlation between impact count and preseason-to-postseason changes in FA and MD. These are presented in Figure 7.

**Figure 7.**
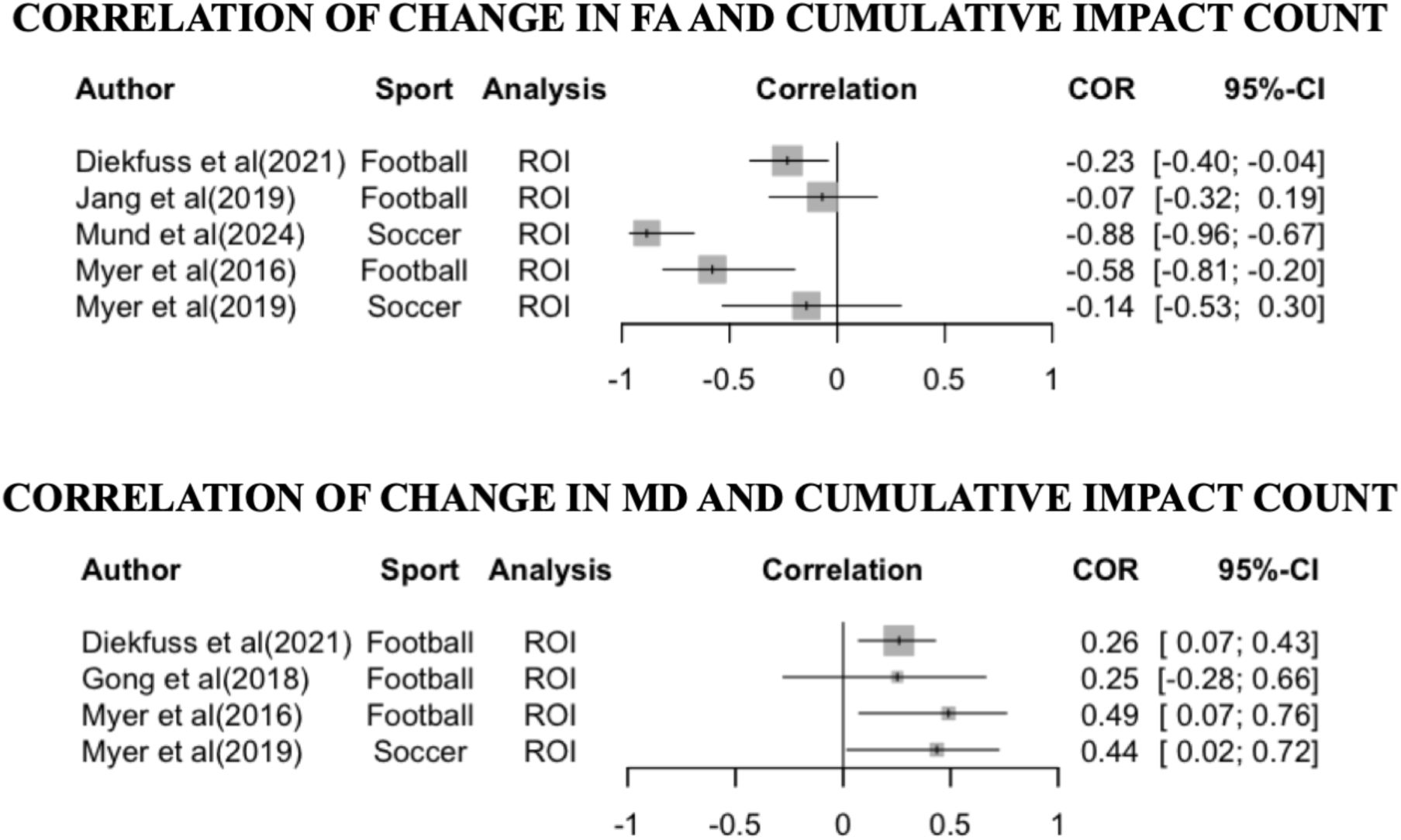
Forest plot of correlation between total number of impacts and longitudinal FA and MD changes. COR (Correlation coefficient), 95%-CI (Confidence Interval), t^2^ (Heterogeneity Variance).

For FA, studies reporting the correlation of cumulative head impact and FA changes reveal a negative correlation. One large negative correlation was observed (d=-0.88)[84], one moderate(d=-0.58)[48], and the remaining are small. On the other hand, MD and cumulative head impact count were reported to be positively correlated with small effect size in four studies.

### Quality ratings

27 of 57 studies were rated high quality, 20 medium, and 10 low. Mean weighted quality score was 89.17 (±8.43). High quality studies were those that reported inclusion/exclusion criteria, method for handling within-in study mTBI diagnosis, and where DTI results were reported accurately including non-significant findings. Among studies with low scores, primary limitations were related to DTI acquisition, processing, and measure used in the study particularly in the DTI protocol utilised and the failure to report non-significant findings in the study. In addition, the majority of studies achieved low in attrition rate of longitudinal subjects. Quality ratings are stated in Table 6, while individual study rating can be found in supplementary material E.

**Table 6.** Quality-assurance-weighted scores rating after consensus.

| Weighted Score (%) | Study | Rating of Weighted Scores |
| --- | --- | --- |
| 100 | Holcomb et al (2021) | H |
| 100 | Kuzminski et al (2018) | H |
| 100 | McAllister et al (2013) | H |
| 100 | Merz et al (2020) | H |
| 100 | Muftuler et al (2020) | H |
| 100 | Wright et al (2021) | H |
| 100 | Diekfuss et al. (2021) | H |
| 100 | Tayebi et al. (2024) | H |
| 100 | Bazarian et al. (2024) | H |
| 94 | Asselin et al (2020) | H |
| 94 | Barber Fross et al (2019) | H |
| 94 | Bazarian et al (2012) | H |
| 94 | Bazarian et al (2014) | H |
| 94 | Davenport et al (2014) | H |
| 94 | Davenport et al (2016) | H |
| 94 | Lancaster et al (2016) | H |
| 94 | Lancaster et al (2018) | H |
| 94 | Manning et al (2020) | H |
| 94 | Murdaugh et al (2018) | H |
| 94 | Myer et al (2019) | H |
| 94 | Puvvada et al (2021) | H |
| 94 | Schranz et al (2018) | H |
| 94 | Strauss et al (2021) | H |
| 94 | Zimmerman et al (2021) | H |
| 94 | Mund et al. (2024) | H |
| 94 | Nilsson et al. (2019) | H |
| 94 | Diekfuss et al. (2021) | H |
| 89 | Champagne et al (2019) | M |
| 89 | Goubran et al(2023) | M |
| 89 | Tayebi et al (2024) | M |
| 89 | Wu et al (2020) | M |
| 88 | Chen et al (2022) | M |
| 88 | Chrisman et al (2016) | M |
| 88 | DiCesare et al (2020) | M |
| 88 | Gong et al (2018) | M |
| 88 | Merchant-Borna et al (2016) | M |
| 88 | Myer et al (2016) - A | M |
| 88 | Myer et al (2016) -B | M |
| 88 | Slobonuv et al (2017) | M |
| 88 | Sollmann et al (2018) | M |
| 88 | Yuan et al (2018) | M |
| 83 | Chun et al (2015) | M |
| 83 | Hunter et al (2019) | M |
| 83 | Jang et al (2019) | M |
| 83 | Mayinger et al (2018) | M |
| 83 | Miller et al (2022) | M |
| 83 | Kelley et al. (2021) | M |
| 82 | Bahrami et al (2016) | L |
| 82 | Dudley et al (2020) | L |
| 82 | Koerte et al (2012) | L |
| 78 | Churchill et al (2020) | L |
| 78 | Brett et al. (2021) | L |
| 76 | Lao et al (2015) | L |
| 76 | Marchi et al (2013) | L |
| 76 | Saghafi et al (2018) | L |
| 67 | Chen et al (2023) | L |
| 60 | Gajawelli et al (2013) | L |

H, high( $\geq Q3=94$ ); M, medium ( $Q1=83 \leq X < Q3=94$ ); L, low ( $< Q1=83$ )

## DISCUSSION

The aim of this systematic review is to assess the current research landscape on the effects of repetitive subclinical HAE exposure on the brains of contact or collision sports athletes, with a particular focus on the various HAE exposure techniques that studies have utilised and how these are correlated with pre- and post-season DTI changes. This is with emphasis on highlighting the filtering or weighting techniques studies have developed to elucidate better correlation of HAE exposure with DTI changes and the common pitfalls encountered by studies utilising these.

In addition, although this review focuses on studies examining HAE, some also included non-contact athlete or non-athletic controls or individuals diagnosed with sports-related mTBI. Findings highlighting differences between these groups will also be discussed. Overall, here we present a broader review discussing studies using DTI and kinematic measures to study the effects of subclinical HAE exposure and further including more synthesised studies.

### Diffusion tensor imaging findings

The impact of subclinical HAE on WM integrity, as measured by DTI, remains inconclusive due to substantial methodological variability across studies [33]. Across studies, FA was the most commonly investigated metric, showing both increase and decreases among contact or collision sport athletes. A similar heterogeneous trend in MD, AD, and RD was also revealed. While some studies report no significant changes, this overall inconsistency highlights a lack of convergence in the current literature. Quantitative synthesis also revealed considerable heterogeneity across included studies, mostly in the direction of observed changes, and reporting methods.

A major contributor to this variability is methodological heterogeneity across studies. As identified in our review, differences in imaging protocols, including field strength, b-values, and number of diffusion directions, as well as variation in preprocessing pipelines and analytical strategies, limit comparability. Although prior reviews have consistently highlighted variability in imaging protocols, pre-processing pipelines, and analytical strategies, yet this heterogeneity persists in more recent investigations[12,31]. These inconsistencies underscore the urgent need for consensus on standardised imaging protocols and acquisition parameters, as previously recommended.^31^. In addition, greater uniformity in analytical approaches should be considered to enhance comparability across studies.^12^ A combined strategy—such as an initial voxel-wise whole-brain analysis followed by targeted ROI-based investigation—may improve sensitivity while maintaining anatomical specificity. In addition, transparent reporting of nonsignificant findings is also essential to reduce publication bias and improve the interpretability of the evidence base. Collectively, methodological harmonisation and transparent reporting are critical to advancing understanding of the relationship between subclinical HAE and WM integrity.

Beyond methodological factors, variability in DTI findings may also reflect distinct and time-dependent neurobiological responses to injury. Decreased diffusivity has been associated with cytotoxic oedema and inflammation, while increased diffusivity is associated with demyelination and axonal loss [92–94]. In addition, microstructural alteration may begin with inflammation associated with increased FA, and progress to axonal injury associated with decreased FA. Both increase and decrease may simultaneously happen. Bazarian et al.[51] found both increases and decreases in FA and MD across the whole WM of each subject when comparing post-season scan to baseline measures. Therefore, in the context of timing, opposing trends in studies is partly confounded by a complex simultaneous process of injury progression[76].

At the group level, these temporal dynamics may obscure true effects, as athletes in early-stage responses (e.g., increased FA) may counterbalance those in later-stage injury (e.g., decreased FA). Unlike mTBI in which there is a single incident as reference of this temporal progression, exposure to subclinical HAE represent a cumulative effect[32]. Moreover, some diffusion changes—such as decreased RD—may reflect improved WM organisation associated with physical activity rather than injury – sports participation in this case[95]. Therefore, similar DTI patterns may represent either injury-related pathology or adaptive neuroplasticity, depending on context and timing. Additional factors—including cumulative exposure, temporal distribution of impacts, biomechanical characteristics (e.g., linear versus rotational acceleration), timing of imaging, and protective equipment—further contribute to the observed variability and should be carefully considered and reported in future studies.

Despite heterogeneity in the directionality of diffusion changes, studies consistently identified WM tracts that are vulnerable to subclinical HAE, mirroring those affected in mTBI[33]. The corpus callosum and superior longitudinal fasciculus were identified to be prominently affected, highlighting their vulnerability to subclinical HAE. These WM tracts play a crucial role in neurobehavioral operations and cognitive capacities[96]. The superior longitudinal fasciculus, for instance, is responsible for integration of sensory information and motor planning, supporting visuospatial attention and complex motor behaviours[97]. This raises concerns, especially among adolescents, as these WM tracts are still developing [70], and emerging evidence suggests an association between WM disruption and long-lasting emotional and behavioural problems.[98]

Current evidence suggests that repetitive subclinical HAE result in intermediate WM alterations between mTBI-diagnosed or concussed athletes and non-contact controls. Bazarian et al. demonstrated that athletes exposed to subclinical HAE exhibited FA and MD changes that were three times greater than those of controls but less pronounced than those of concussed or mTBI-diagnosed athletes [91]. Goubran et al. similarly found intermediate WM changes in football players relative to volleyball players, suggesting a spectrum of WM integrity influenced by subclinical HAE exposure[75]. Even though inconclusive, these studies imply that subclical HAE, though milder than mTBI, still lead to measurable WM changes [19,75,80,99,100]. Whether repetitive subclinical HAE ultimately results in brain alterations comparable to those seen after a single mTBI remains an important and evolving area of investigation.

In addition, several studies suggest a dose-dependent relationship, indicating that higher cumulative HAE negatively affect WM integrity and cognitive function. Strauss et al. showed that athletes with low exposure to HAE had beneficial neural adaptations, while those with high exposure lacked such advantages [88]. Additionally, Holcomb et al. identified significant FA changes in athletes experiencing high cumulative strain compared to controls, which were absent in those with lower exposure [64]. These findings support the notion that HAE may exist along a continuum, in which limited exposure could be associated with adaptive neuroplastic responses, while excessive cumulative burden may exceed the brain’s compensatory capacity and lead to microstructural disruption. Importantly, this raises critical questions regarding potential exposure thresholds at which repetitive HAE transition from adaptive to deleterious. Establishing empirically derived exposure limits—integrating impact frequency, magnitude, and individual susceptibility—remains essential for informing athlete monitoring, return-to-play policies, and long-term risk mitigation strategies.

A critical factor contributing to inconsistent findings is substantial inter-individual variability in WM responses to repetitive HAE. Studies relying on group-level analyses may overlook subject-specific patterns, potentially leading to conflicting results. Chun et al. demonstrated that different collision intensities within teams led to opposing FA trends, emphasising the variability in injury mechanisms and responses [54]. Asselin et al. employed the SPREAD algorithm, highlighting significant but spatially varied FA decreases at an individual level, which group-level analysis could not detect [66]. Similarly, Bazarian et al. implemented wild bootstrapping to reveal individual-level changes obscured by traditional analyses [91]. Collectively, these findings highlight the importance of personalised or subject-level analytical frameworks to more accurately characterise WM alterations associated with HAE.

Variations in DTI metrics reflect distinct underlying pathological processes. Many studies report a pattern of decreased FA accompanied by increased MD, AD, or RD, indicative of demyelination and axonal injury [36,47,75,101]. Conversely, increased FA coupled with decreased diffusivities suggests inflammation or transient axonal changes [61,73,86]. Additionally, Gong et al. reported microstructural changes in cortical and deep gray matter, showing increased MD and mean kurtosis in the thalamus and putamen, and region-specific cortical changes [74]. Mixed evidence exists regarding WM recovery following rest periods from contact sports. Bazarian et al. reported persistent group-level changes even after six months of rest, though individual athletes varied substantially in recovery trajectories [91]. Taken together, these findings emphasise that interpretation of DTI changes must account for injury timing, chronicity, cumulative exposure, and pronounced inter-individual variability.

### HAE metrics

Various metrics have been employed to quantify HAE, including impact counts, linear and rotational accelerations measured by sensors, head injury measures (HIC, GSI, HITsp), and FE analysis-derived strain measures. Schneider et al. demonstrated that greater exposure to HAE correlates with more significant WM changes [32]. Consistent with this, our quantitative synthesis identified negative correlations between total impact count and FA, alongside positive correlations with MD, suggesting microstructural disruption. However, these only revealed small degree of correlation. This is potentially due to total impact counts not fully capturing variations in impact severity, magnitude, or direction [76,102,103]. Two athletes with similar impact frequencies may experience substantially different biomechanical burdens depending on these factors. Consequently, reliance on frequency-based metrics may oversimplify exposure characterisation and partially explain inconsistencies across studies.

It should be noted that among the studies included in this review that used impact count as a measure of subclinical HAE exposure, only a few applied additional magnitude-based thresholding. Diekfuss et al.[62] quantified impact count thresholded at 20g to 150g in intervals of 10g (e.g. >20g, >30g). Significant interactions between season-long DTI changes and impact count were observed only at higher thresholds (110–140 g). These findings suggest that white matter alterations were primarily driven by higher-magnitude impacts rather than cumulative exposure alone. Importantly, this highlights that the biological relevance of HAE may be severity-dependent. However, the optimal threshold at which impacts become meaningfully associated with microstructural change remains unclear and is an active area of investigation.

Furthermore, studies indicate that cumulative, time-weighted, and risk-weighted metrics correlate more strongly with FA than simpler measures such as peak or mean accelerations. These weighted metrics account for factors such as recovery intervals, the risk associated with individual impacts, and cumulative exposure effects, highlighting their importance for accurately assessing HAE exposure [36,57]. Linear acceleration of the head has been associated with a transient gradient in intracranial pressure, while rotational acceleration is associated with strain[90]. There remains an ongoing debate over whether linear or rotational acceleration is the primary driver of microstructural alteration. While these measures have been commonly used in HAE studies separately, particularly those included in this review, recent investigations have highlighted the importance of accounting for both in a single combined probability metric[36,72]. In addition, risk-weighting has been developed to account for the risk of concussion or injury for each HAE[104]. Incorporating these, Davenport et al. has revealed better correlation of risk-weighting combined probability with DTI metric changes than risk-weighted linear or rotational acceleration. However, it should be noted that these metrics have been developed from data on football players characterised by a lack of impact, predominantly rotational acceleration[90]. Further study is warranted to test its applicability to other sports or to develop a sport-specific risk curve.

On the other hand, metrics were also developed to account for acceleration, duration, and location of each impact[105]. These include the HIC, GSI, and HITsp. Merchant-Borna has shown a stronger correlation of HIC and GSI than linear and rotational acceleration[81]. However, included studies in this review that tested and found a correlation of these measures with DTI changes had football or hockey player cohorts[51,66,80,81]. This is partly due to the prevalence of these metrics in helmet-mounted accelerometer studies. To which studies have been concerned regarding helmet slippage when the helmet is not snug to the skull, causing differential motion between the head and helmet[81,106]. There remains a lack of data on the applicability of these metrics in sports where HAE exposure monitoring is feasible only with mouthguard- or patch-based devices (*e.g.,* rugby).

In terms of time-weighting, Asselin et al. found a significant correlation between time-weighted HAE and subject-specific WM changes[66]. Specifically, time-weighted approaches like TBH and TUA improve sensitivity in detecting neuronal vulnerability relative to impact timing [81,107,108]. TBH accounts for the number, magnitude, and timing of prior impacts, suggesting that even sub-threshold impacts can cause axonal injury if they occur within a short time window [107]. TUA considers the time between impact and DTI assessment, with injury less likely to be observable as this interval increases [81,108]. Integrating these concepts, a combined time-weighting incorporating TUA+TBH was applied which revealed improved correlation with DTI changes[81]. This reveals the importance of accounting for time when correlating HAE exposure with microstructural alterations. From a subject-specific perspective, the efficacy of these models is often challenged by the heterogeneous timing of HAE accumulation; while some athletes sustain heavy exposure early in a season, others may experience a surge in impacts near the end. This variability is further compounded by the non-linear, concurrent processes of axonal injury and physiological repair within the same individual[76]. Consequently, a single post-season snapshot may capture a ’net zero’ mean change that masks significant underlying disturbances. This highlight the critical necessity of further optimising multi-dimensional weighting techniques—specifically those that integrate TBH and TUA—to resolve the temporal complexities of white matter alterations and improve the quantitative precision of HAE assessments.

Up to this point, numerous metrics have been widely used. However, these consider only the peak values of kinematic response[83]. They do not account for region-specific responses, which limits their ability to distangle the regional variation of injury-causing kinematics for a given HAE event. The use of FE modelling reveals promising results in correlating an injury and the regional tissue changes. In this review, FE-derived strain metrics have shown superior sensitivity in correlating HAE with WM alterations compared to acceleration measures. Miller et al. identified strain metrics as the best-performing indicators of WM changes, surpassing traditional impact metrics (*e.g.*, RWE) [83]. These strain metrics account for regional differences in brain stiffness, providing valuable insights into specific WM tract vulnerabilities [57]. Additionally, region-of-interest-based analyses have identified specific correlations between HAE and DTI alterations in regions such as the corpus callosum and superior longitudinal fasciculus [60,78,82]. With regionalisation of strain, understanding the relation between HAE and DTI changes as an effect of injury can be further understood. While strain measures reveal superior performance, existing studies correlating these have used a generic head model with and an assumption that the brain is homogenous and isotropic[64]. Therefore, future direction for this measure involves integration of white matter fibre orientations, white matter and grey matter material differences, and utilising subject-specific head models to better characterise strain propagations.

Fundamental to these measures is the accurate quantification of HAE exposure during sports[73]. To this, kinematic sensors as presented in this review have been used. A common approach to reduce false positives involves setting an arbitrary peak acceleration threshold. However, traditional thresholding techniques often fail to differentiate true impacts from artifacts. Jang et al. demonstrated a correlation between cumulative impacts above 20g and DTI changes, but false positives remained problematic [76,109]. This is partially due to the limitations of thresholding in differentiating real HAE from athletic activities, such as the removal of the instrumented helmet, which can still cause acceleration. Recent machine learning-based verification methods have significantly improved accuracy, reducing false positives and enhancing correlations with DTI metrics [73]. Such advanced analytical techniques are essential for reliable data interpretation. Such findings support the need for advanced computational modelling and standardised, sport-specific metrics to better characterise cumulative HAE exposure and its relationship to neuroimaging outcomes.

Given the substantial variability in metrics and methods, current quantitative findings should be interpreted cautiously. Future research should focus on validating standardised, sport-specific approaches that integrate filtering, weighting, and modelling techniques to improve accuracy, enhance mechanistic understanding of WM injury, and inform strategies for athlete safety.

### Other MRI modalities

While most research has focused only on DTI, a few studies have employed other MRI modalities to better understand WM changes from repetitive HAE. Notably, three studies using DKI reported significant alterations that were not always captured by DTI alone [6,74,110]. For example, Davenport et al., in two studies involving the same cohort, found both DTI and DKI changes, suggesting DKI may offer increased sensitivity to HAE [72,103].

Other imaging techniques such as functional MRI, Magnetic Resonance Spectroscopy, and Arterial Spin Labeling have also revealed meaningful brain alterations [10,53,86]. While the detailed analysis of these methods is beyond the scope of this review, integrating multi-modal MRI approaches may provide a more comprehensive picture of brain response to repetitive HAE and support findings from DTI studies.

### Confounding variables and future direction

Building on the methodological and biological variability discussed above, several key confounding factors further complicate interpretation, including age, mTBI history, clinical and physiological factors, and study methodologies. Age-specific differences in brain maturation and susceptibility to HAE remain uncertain, as evidenced by limited WM changes observed in youth athletes compared to more pronounced alterations in high school athletes. However, small sample sizes and variations in timing and intensity of exposure complicate the interpretation of these results, suggesting the need for careful age considerations in future research.

Sex-related differences also affect WM responses to HAEs. Studies indicate males and females may experience distinct WM changes, potentially due to differences in physiological and hormonal factors and variability in impact frequency or severity. Additionally, prior mTBI significantly influences susceptibility to subsequent HAE and WM alterations, underscoring the importance of carefully documenting and controlling for mTBI history. Variability in the management of mTBI subjects across studies further complicates interpretation.

Methodological differences, such as voxel-based versus ROI-based analyses, group averaging, and the inclusion of control groups, influence results. Future studies should adopt comprehensive approaches incorporating both voxel-based and ROI-based analyses, subject-specific approaches, standardised control groups - particularly non-contact sports athletes-, and better characterisation of clinical variables to enhance understanding and reduce variability.

In terms of HAE quantification, studies should adopt combined weighting techniques (*e.g.,* TUA, TBH, RWE) to better resolve timing and HAE accumulation relative to scanning time, the time between HAEs, and the risk. Furthermore, validation of risk curves and time filtering should be made in sporting cohorts apart from football and hockey to tests its generalisability. The use of FE in the regionalisation of injury also holds promise to better map HAE exposure to observed DTI changes or injury. Overall, while multiple metrics have been established to quantify HAE exposure, extensive validation and utilisation are still warranted.

### Strengths and Limitations

Current findings provide evidence of the structural changes over a single season of contact sports participation. This review underscores key methodological considerations and offer direction for future HAE studies. A modified NIH quality rating tool, adapted from Lees et al. was used to assess the risk of bias in included studies and may serve as a framework for future evaluations.

This review has several limitations. First, potential publication bias exists, as studies with non-significant findings may be underreported. Second, only longitudinal studies on non-mTBI diagnosed or non-concussed contact sport athletes (and those examining mTBI-diagnosed or concussed contact sport athletes as a comparison group) were included. While this approach provided useful insights, it may have introduced heterogeneity. To mitigate this, additional inclusion criteria were applied to studies in the quantitative synthesis.

Third, reported effect sizes may be underestimated due to incomplete statistical data. Although this review acknowledged findings from other MRI modalities, the analysis focused solely on DTI, leaving room for future studies to explore multi-modal imaging and its relationship with DTI. Fourth, the review did not assess correlation between DTI parameters and clinical symptoms, such as cognitive function or neuropsychological outcomes. Investigating these relationships would improve our understanding of the clinical relevance of DTI findings.

Finally, while a tailored version of Lees (2021) quality tool was employed to assess study quality, this may have impacted ratings. It is important to note that exclusion of a study based on these criteria does not imply poor quality, but rather that the study did not fully meet the specific requirements for inclusion.

### Study implications

Overall, sports participation offers physical, psychological, and social benefits[1], and improves cardiovascular and metabolic health[2]. However, observed longitudinal DTI changes associated with underlying pathophysiology emphasise concerns about athletes’ short- and long-term brain health following repetitive subclinical HAE.

While longitudinal DTI alterations are widely observed following repetitive HAE, the underlying dose-dependent mechanisms remain poorly characterised. By synthesising evidence of how cumulative HAE exposure correlates with microstructural disruption, this review warrants the identification of a threshold at which mechanical loading initiates subtle WM disorganisation. Our synthesis reveals emerging evidence of a dose-dependent relationship, where higher cumulative HAE exposure correlates with greater microstructural disruption, potentially counteracting the broader benefits of athletic participation. These findings support implementing structured “no-collision” recovery periods to promote neural healing and provide a foundation for evidence-based HAE exposure monitoring, injury prevention, and guided athletic rehabilitation.

## CONCLUSION

This systematic review and quantitative synthesis provide evidence of longitudinal WM alterations - inconsistent FA, MD, AD, and RD findings - associated with participation in collision sport, as detected by DTI. The findings demonstrate significant heterogeneity across studies, likely influenced by variations in study design, sample characteristics, imaging protocols, and analysis techniques. Furthermore, inconsistencies suggest that individual exposure factors such as HAE history, recovery time, and methodological differences in estimating exposure may contribute to these variations.

Quantitative synthesis of a subset of studies that assessed the relationship between HAE exposure and WM alterations yielded consistent effects. This suggests a dose-dependent relationship between subclinical HAE exposure and WM integrity indicating that higher cumulative HAE is associated with greater microstructural disruption. Taken together, these observations underscore the importance of adjusting for individualised HAE exposure profiles when interpreting longitudinal WM changes in collision sport athletes.

Future research should focus on standardising imaging protocols and HAE quantification methods, integrating multimodal imaging approaches, and improving subject-specific analysis techniques to enhance the accuracy and interpretability of findings. The evidence highlights the potential impact of HAE on brain health, underscoring the need for continued monitoring and refined methodologies to assess long-term risks in collision sport athletes. Further work is needed to understand whether the implementation of structured breaks from HAE exposure facilitates neural recovery. Collectively, these recommendations may lead to evidence-based injury prevention and rehabilitation guidelines to mitigate potential neurological sequelae associated with HAE exposure

## Data Availability

Data sharing not applicable to this article as no datasets were generated or analysed during the current study.

## ACKNOWLEDGEMENT

Supported by Royal Society of New Zealand Marsden Fund, Neurological Foundation, Kanoa – Regional Economic Development & Investment Unit, New Zealand; Trust Tairāwhiti, Catalyst Strategic Fund from Government Funding administered by the New Zealand Ministry of Business Innovation and Employment, Fred Lewis Enterprise Foundation, the Hugh Green Foundation, JN & HB Williams Foundation, New Zealand Health Research Council, Anonymous Donation, and Te Tītioki Mataora. CS is supported by the Department of Science and Technology – Science Education Institute, Foreign Graduate Scholarship and the University of the Immaculate Conception, Davao City, Philippines. MT and EK is supported by a Hugh Green Foundation Senior Research Fellowship. JM is supported by a Philip Wrightson Fellowship. WS is supported by a Vision Research Foundation Senior Research Fellowship.

## AUTHOR DISCLOSURE STATEMENT

No competing financial interest to be declared by the authors.

### Summary of Studies

• Asselin et al. (2020)[66] The authors conducted a secondary analysis of data from contact sport athletes (N=28) in a longitudinal study design to determine the correlation of helmet data and longitudinal changes in DTI using the SPatial Regression Analysis of DTI (SPREAD). Prominently detected injured regions were associated with decreased FA values, yet these regions occupied the whole WM skeleton, suggesting a subject-specific injury pattern.

• Bahrami et al.(2016)[70] The authors examined the effect of HAE measured using HITS resulting from a single season of youth football in WM tract-specific DTI metrics. This was conducted in a cohort of male football players (N=25). The study found a significant correlation between the decrease in FA of the SLF terminal and the whole left IFOF, its core, and terminals with Risk-weighted exposure.

• Barber Foss et al. (2019)[45] The authors sought to determine preseason to postseason changes in the WM integrity measured using diffusion tensor imaging from HAE exposure in a cohort of High School Football Players (N=21) and Youth Football Players (N=12). High school players exhibited a reduction in MD, AD, and RD in widespread WM areas, while the youth players only had reduced AD to a more limited extent. Hence, it does not confirm that younger children are susceptible to the effects of HAE.

• Bazarian et al. (2012)[71] The authors performed a study on athletes engaged in football and hockey (N=9) and controls (N=6) over a season to investigate the ability of wild bootstrapping to detect subject-specific WM changes in WM and its association with self-reported HAE count and cognitive scores. Wild bootstrapping detected significantly changed WM in a subject-specific pre- and post-season comparison, revealing the percentage of WM with a significant change to be three times higher in athletes subjected to HAE than controls.

• Bazarian et al. (2014)[51]The authors conducted diffused tensor imaging in a cohort of collegiate football players (N=10) with helmet impact sensor measures before and after a single season and after six months of no-contact rest. This was made to quantify HAE with WM changes, including relevance in terms of cognitive function and clinical tests. Among football players, FA and MD changed from pre- to post-season, with most differences persisting after six months of no-contact rest. The percentage of voxels with decreased FA was correlated with HAE measures and changes in serum ApoA1 and S100B autoantibodies.

• Bazarian et al.(2024)[59] The authors tested whether blood-based biomarker changes are correlated with HAE exposure and DTI changes. A significant decrease in FA of the fornix was observed. Changes in blood-based biomarkers also showed correlation with FA reduction in the fornix and medial meniscus, and MD of the fornix.

• Brett et al. (2021)[89] The authors in a cohort of high school and collegiate athletes (N=121) assessed four times tested the association of years of HAE exposure and prior mTBI with diffusion metrics of WM organisation including volume as well as quantitative susceptibility mapping. Results reveal the association of cumulative years of HAE exposure with elevated FA and decreased RD.

• Champagne et al. (2019)[57] The authors combined diffusion tensor imaging, HAE exposure measure, and amplified magnetic resonance imaging to observe region-specific microstructural changes in the corpus callosum upon exposure to HAE in a longitudinal observational study of football players (N=33) observed at pre-season, mid-season, and post-season. Their approach revealed specific differences in strain along the WM tract, which suggests a possible relationship between the WM tissue properties and its integrity.

• Chen et al. (2022)[111]The authors studied athletes with sports-related mTBI (N=24), athletes with repetitive HAE (N=26), and non-contact sport athletes (N=28) in 4 time periods to investigate WM microstructural differences with and without sports-related mTBI. A significantly increased axial kurtosis in athletes with SRC compared with controls was observed, which also manifested when repetitive HAE athletes were compared with controls. The extent of WM regions manifesting difference decreased across time points yet remained present primarily in the Corpus Callosum.

• Chen et al. (2023)[67] The authors in a cohort of collegiate sport athletes (N=36) and non-contact sport controls (N=45) created a machine learning algorithm that classifies these groups based on diffusion metrics. MD, mean kurtosis, and extra-axonal compartment diffusion revealed to be the most influential metric for classification.

• Chrisman et al. (2016)[58]The authors conducted a prospective cohort study measuring the frequency and magnitude of HAE in middle school-age soccer players (N=17) over a weekend tournament. They tested the association of HAE measures with cognitive scores, DTI metric changes, and advanced neuroimaging metrics. No significant associations were found between HAE exposure, neuropsychological testing, and advanced neuroimaging indices after correction for multiple comparisons.

• Chun et al.(2022)[54] The authors longitudinally evaluated diffusion-weighted images and collision event monitoring in a cohort of high school athletes (N=34) participating in a single American football season. The study revealed a correlation between head collision events and changes in DTI measures in the Left SCR and right SFOF for one group of athletes and right RLIC for the other. However, FA increased in the first group, while FA decreased in the second group, suggesting varying axonal insults and leaving an open question for clinical relevance.

• Churchill et al.(2020)[112] The authors determined subject-specific longitudinal changes in a cohort of university-level athletes (N=123) scanned pre-season. Of this group, concussed athletes (N=12) were scanned at symptomatic injury and returned to play, while a subgroup of non-concussed athletes (N=44) were scanned post-season. The study highlighted the impact of the use of pre-injury baseline data in the study of mTBI-related brain abnormalities in a subject-specific focus, as well as the necessity of both longitudinal and cross-sectional comparisons between cases and controls.

• Davenport et al. (2014)[72]The authors determined whether the cumulative effect of HAE, particularly Risk Weighted Linear Acceleration, Rotational Acceleration, and Combined Probability from high school football season in athletes (N=24), caused MRI metric changes without mTBI diagnosis. Their analysis revealed a statistically significant association between RWECP and FA, while a secondary analysis revealed an association between RWELinear and RWECP and FA, MD, CP, CL, and CS. In addition, a strong correlation was revealed between DTI measures and changes in the Verbal Memory subscore of ImPACT. Thus demonstrating the effect of a single season of playing football on the athlete’s brain without a clinical diagnosis of mTBI.

• Davenport et al. (2016)[68] The authors determined whether the cumulative effect of HAE in a season of high school football in athletes (N=14) is associated with changes in diffusion kurtosis metrics without a clinical diagnosis of mTBI. Findings reveal a significant relationship between RWECP and Mean Kurtosis, while secondary analysis demonstrated a significant relationship between RWECP and DKI-derived metrics after covariate adjustment. Compared with DTI, diffusion kurtosis metrics they explained more variance, suggesting that it may be more sensitive to HAE.

• DiCesare et al.(2020)[73] The authors examined whether data-driven filtering of HAE exposure using machine learning classification produces a more accurate quantification of exposure to HAE and reveals a more pronounced relationship with longitudinal brain changes. This was performed in a longitudinal study of female high school soccer players (N=22) for one season. Findings reveal that the ML approach achieved higher accuracy in filtering HAE compared with a threshold and heuristic approaches while also revealing a significant association with longitudinal WM changes.

• Diekfuss et al.(2021)[62] The authors investigated whether internal jugular vein compression via a neck collar modulated the relationship between HAE exposure and pre- to post-season DTI changes in male high school football athletes (N=284). Associations between high-magnitude head impacts and longitudinal DTI changes varied in direction between collar and non-collar groups. Athletes wearing the collar showed partial preservation of DTI metrics over the season, suggesting a mechanistic protective effect of JVC on white matter.

• Diekfuss et al.(2021)[63] The authors tested whether wearing newer, higher-ranked football helmets versus old helmets affect seasonal alteration in DTI metrics. This is in a cohort of high school athletes (N=54 for newer; N=62 for old). Subjects wearing the old helmets displaced reduction in MD, AD, and RD. On the other hand, subjects wearing newer helmets revealed increase in AD.

• Dudley et al.(2020)[53] The authors evaluated the efficacy of a jugular vein compression collar for preserving the functional and structural measures of brain network organisation in a single season of competitive play among a cohort of female high school soccer players (N=128). Non-collar-wearing athletes exposed to repetitive HAE exhibited significantly increased rs-fMRI-derived global clustering coefficient and DTI-derived modularity compared to collar-wearing athletes. Thus, this adds to the body of literature reporting the alteration of WM integrity and measures of brain structure after participation in contact sports.

• Gajawelli et al.(2013)[19] The authors determined if differences in diffusion tensor imaging between contact sport athletes (N=11) and non-contact sport athletes (N=13) caused by repeated physical impact on the brain were evident, including pre- and post-season differences. Findings reveal significant differences when comparing contact sport athletes and non-contact sport athletes and contrasting pre- to post-season scans of contact sport athletes in the IFOF, SCR, PCR, and sCC FA values. Small clusters of difference are also found in the gCC and bCC.

• Gong et al.(2018) The authors utilised Diffusion Kurtosis Imaging and Quantitative Susceptibility Mapping to probe microstructural changes in the gray matter in a cohort of male football players (N=16) over a single season. Changes were tested for correlation with HAE. A decrease in MK and an increase in MD were observed in the Thalamus and Putamen. DKI metrics were correlated with HAE metrics, revealing the potential of DKI to yield valuable biomarkers for evaluating the severity of brain injuries.

• (Goubran et al., 2023)[74] The authors investigated longitudinal changes in the brain’s microstructure measured using diffusion MRI and automated fibre quantification among collegiate contact(N=49) and non-contact(24) athletes. A longitudinal increase in FA and AWF and a decrease in RD, MD, and ODI was found in non-contact athletes, which were relatively absent in contact sport athletes. This suggests that normative changes in diffusion metrics associated with healthy brain development may be altered and diminished in contact with sports athletes.

• Holcomb et al.(2021)[64] The authors assessed the relationship between WM diffusion changes and finite element strain measures in a cohort of non-concussed youth football players (N=102). In addition, the authors determined the association between strain intensity (high or low) exposure and DTI metrics. A positive linear relationship in percentage change of FA and cumulative maximum principal strain one time strain rate was identified in the CgC, Fx, IC, EC, CC, CR, CST, CP, SLF, and right SFOF. In addition, the posthoc analysis revealed that strain-based metrics significantly explained more variance in the FA change than several HAE alone.

• Hunter et al.(2019)[113] The authors examined the potential effect of BDNF Val66Met polymorphism in the association of soccer heading and WM microstructure measured by diffusion tensor imaging in a cohort of soccer players (N=312). Findings revealed a significant interaction of 12-month heading *BDNF Val66Met Genotype on the presence of low RD.

• Jang et al.(2019)[76] The study explored whether counts of head acceleration events are associated with changes in traditional diffusion-based measures, particularly FA and MD, in a cohort of football players (N=61) compared with non-collision sporting peers (N=15). The spatial extent of the football athlete brain exhibiting DTI changes was associated with cumulative HAE having peak translational acceleration of more than 20g. Thus adding to evidence that HAEs cause low-level neurotrauma, with prolonged exposure producing a more significant accumulation of neural damage.

• Kelley et al. (2021)[69] The authors aimed to characterise changes in HAE across multiple football season and test its correlation with DTI changes. This was tested in a cohort of youth football athletes (N=47; N=19 with imaging data) with multiple consecutive seasons of HAE. Increases and decreases in HAE exposure were observed from one season to another, and changes in the number of practice impacts and in HAE exposure between seasons correlated with changes in the number of abnormal voxels.

• Koerte et al.(2012)[77] The authors investigated the effect of repetitive HAE on WM integrity using diffusion tensor imaging sustained by players (N=17) in a Canadian Interuniversity Sports (CIS) ice hockey season. TBSS revealed an increase in trace, radial diffusivity (RD), and axial diffusivity (AD) from pre- to postseason localised in the right precentral region, right corona radiata, and the anterior and posterior limb of the internal capsule.

• Kuzminski et al.(2018)[78] The authors conducted a longitudinal analysis of football athletes (N=17) using diffusion tensor imaging to assess the WM changes in high school football season and correlate DTI changes with helmet accelerometer and neurocognitive data. The voxel-wise analysis did not reveal significant change across the season. However, ROI analysis showed a significant decrease in FA in the Fornix-stria Terminalis and Cingulum Hippocampus, which found an association with impact frequency.

• Lancaster et al (2016)[114] The authors characterised WM changes within 24 hours and 8 days of mTBI using DTI and DKI in a cohort of concussed (n=26) and nonconcussed (n=26) athletes. Within 24 hours, decrease in MD, AD, and RD, while an increase in AK was observed in the concussed group. This persisted at 8 days post-injury despite symptom resolution. Findings reveal no full physiological recovery a week after injury.

• Lancaster et al.(2018)[115] The authors explored the effects of mTBI, including recovery trajectory in a cohort of concussed football players (N=17) with non-concussed football players (N=20) using diffusion tensor imaging and diffusion kurtosis imaging. TBSS revealed a continued widespread decrease in MD and AD in concussed subjects compared with controls. However, kurtosis indices normalised. Findings add to evidence of the utility of DTI and DKI in assessing mTBI severity, recovery, and long-term effects.

• Lao et al.(2015)[49] The authors conducted a joint analysis of T1 surface-based morphometry and DTI analysis on 3D surface representation of the corpus callosum in a cohort of contact sports players (N=11) in a single season. The combined statistical analysis of the T1-weighted image and DTI increased detection power in detecting differences between pre-and post-season contact sport athletes.

• Manning et al (2020)[79] The authors assessed the longitudinal brain microstructure and function in a cohort of female athletes participation in contact (n=70) and noncontact (n=31) sport athletes. Cross-sectional and longitudinal comparison revealed significant difference and change in diffusion metrics. Specifically, lower FA and higher MD, AD, and RD in contact sport athletes compared to non-contact sport athletes. Hence, demonstrating longitudinal alteration in brain microstructure in healthy, asymptomatic athletes.

• Marchi et al.(2013)[8] The authors tested whether blood-brain barrier disruption (BBBD) and accompanying surge of astrocytic protein S100B in the blood causes immune response through the production of auto-antibodies and determine whether this results in disrupted WM integrity through DTI scans. Transient BBB damage measured using serum S100B was detected only in players exposed to a significant number of HAEs. In addition, serum levels of S100B auto-antibodies were also predictive of abnormalities in DTI indices, which correlate with cognitive change. This correlation between variables supports the notion of the association between BBBD and the potential risk for cognitive changes.

• Mayinger et al.(2018) [52] The authors evaluated longitudinal changes in the diffusion of WM tracts in collegiate football athletes (N=15) in three periods: pre-, post-, and off-season using TBSS. Findings reveal an increase in FA in the left parietal lobe and an increase in trace in the brainstem and left temporal lobe from pre- to post-season. On the other hand, a significant decrease in trace and FA was observed after six months of no-contact rest. This suggests the alteration of WM integrity in contact sport athletes as an effect of a season of contact sport participation, which returns to baseline after a period of no-contact rest, suggesting the benefit of this period for athletes.

• Mcallister et al.(2013)[80] The authors determined whether exposure to repetitive HAEs over a single season affects WM diffusion measures of collegiate contact sport athletes (N=80). Findings reveal a correlation between WM DTI measures of the CC, Amygdala, Cerebellar WM, Hippocampus, and Thalamus and HAE exposure. In addition, the magnitude of MD change in the CC was associated with poorer verbal learning and memory performance.

• Merchant-Borna et al.(2016)[81] The authors developed a novel method for quantifying cumulative HAE by weighting helmet-based measures with time between impact and time upon assessment and testing its association with DTI changes over collegiate football season (N=10). The utilisation of time until assessment and time before hits resulted in an improved correlation of HAE with WM changes, supporting the notion of the deleterious effect of HAEs to sub-clinical brain injury influenced by time-between hits.

• Merz et al.(2020)[82] The authors compared diffusivity metrics in a single season of athletic activity within and between a contact sport athletic cohort (N=75) and non-contact athletes (N=79) and its association with HAE measures and cognitive scores. A Bonferroni correction was applied in the test for significance. Seasonal change in FA and MD did not differ between groups. However, specific to contact sports athletes, a positive association was found between the uncincate fasciculus MD values and season linear acceleration, rotational acceleration, and hit severity. This suggests an influence of various HAE characteristics, including type, severity, and frequency, on specific brain regions.

• Miller et al.(2022)[83] The authors gathered head kinematic data from male Football athletes (N=95) between 2012 and 2017. They tested whether strain-based metrics from finite element analysis of HAE or risk-weighted exposure best discriminate changes in DTI Metrics, particularly FA, MD, CL, CP, and CS. A significant relationship was found between the DTI indices studied and strain measures. On the other hand, significant relationships were found between FA and RWECP and RWELinear, as well as Cs and RWELinear. The area under the receiver operating characteristic curve found that the best-performing metrics were strain measures, suggesting better discriminatory power over kinematic metrics in the study of imaging changes.

• Mund et al. (2024)[84] The authors investigated the effect of heading on WM organisation of high-level adult male football players. While no significant longitudinal change in DTI metrics was observed, a change in FA in the splenium of the corpus callosum correlated with the frequency of long-distance headers.

• Tugan Muftuler et al.(2020)[116] The authors used diffusion tensor imaging and diffusion kurtosis imaging to monitor the effects of SRC on the brain and trajectory of recovery of concussed athletes (N=96) compared with non-concussed athletes (N=82) scanned in 4 times periods. Results of this study indicate that changes in the acute period may be more prolonged compared with clinical recovery and that brain alteration may not be observed in the acute stage yet may appear in later stages of the injury. Thus highlighting a potential disparity between clinical symptoms and pathophysiological recovery from sports-related mTBI evident in neuroimaging findings.

• Murdaugh et al.(2018)[117] The authors investigated the alteration in functional connectivity, WM integrity, and cognitive abilities in a cohort of concussed football players (N=16) and non-concussed football players (N=12) measured acutely after injury and 21 days after baseline measurement. Investigation into the change of DTI indices reveals altered diffusion in the SRC group along the CST and SLF in the acute phase of SRC, which were not evident at follow-up imaging (21 days later).

• Myer et al. (2016) [60] The authors investigated the effect of wearing a collar during HAE exposure on brain microstructure measured using diffusion tensor imaging during football season. The difference in microstructural alterations pre-season and post-season was determined between the collar group (N=21) and the non-collar group (N=21). Collar and non-collar groups experienced similar overall g forces and HAE exposure, yet only in the non-collar group were changes in MD, AD, and RD observed.

• Myer et al.(2016)[48] The authors evaluated the use of mild neck compression applied during HAE exposure in reducing mild neck compression applied during HAE exposure in reducing anatomical biomarkers of brain injury. This was measured in a cohort of rugby players divided into a collar group (N=7) and a non-collar group (N=8). DTI measures, mainly the MD and RD, significantly increased from pre- to mid-season, corresponding to WM microstructure disruption. However, not related to DTI, the study provides a direction towards protecting the brain from sports-related HAE through optimisation of intracranial fluid dynamics.

• Myer et al.(2019)[45] The authors quantified WM alteration in a female high school soccer player cohort (N=46) in pre-, post-, and off-season and determined association with measures of HAE, particular total HAE count, cumulative g-force, and average g-force. Findings reveal significant pre- to post-season WM changes in athletes not wearing the jugular vein compression collar, particularly regarding MD, AD, and RD. A significant correlation was also found between DTI changes and HAE exposure. These differences were resolved at three months of off-season follow-up.

• Nilsson et al. (2019)[85] The authors aimed to assess the effect of repetitive HAE exposure in a cohort of 8-12-year-old collision sport athletes (N=35) longitudinally or in comparison with non-collision sport athletes (N=12). There was no significant group by time interaction in DTI metrics. However, the total magnitude of lateral head impacts revealed positive and moderate association with increase in FA of the left cingulate cortex.

• Puvvada et al.(2021)[36] The authors compared cumulative HAE exposure metrics derived by Risk-weighted Exposure and Time-weighted Exposure with directional changes in DTI metric in a longitudinal study of football players (N=95). The findings of the study reveal that cumulative biomechanical metrics were more associated with an increase in abnormal voxels than comparison with a decrease in abnormal voxels. The study’s results warrant further investigation into the physiological phenomena represented by directional change.

• Saghafi et al.(2018)[50] The authors utilised a convolutional neural network to classify players (N=60) into high and low-risk-weighted exposure measured using helmet-mounted accelerometers with change in pre- to post-season FA maps as predictor variables. The algorithm accurately classified athletes subjected to HAE into high and low-impact exposure. The work adds to evidence of detectable neuroimaging brain changes in athletes after playing a single season of contact sports.

• Schranz et al.(2018)[86] The authors used non-invasive Proton Magnetic Resonance Spectroscopy (MRS) and DTI to monitor changes in prefrontal WM metabolic changes and brain microstructural integrity in female rugby players with and without concussed (N=64). Within the voxel where glutamine changes were observed in MRS, an increase in FA and a decrease in RD were observed in non-concussed athletes, which may indicate neuroinflammation or re-myelination. These findings were not correlated with clinical test scores suggesting brain imaging is more sensitive to brain injury, which raises its potential of aiding in brain injury recovery.

• Slobounov et al.(2017)[10] The authors examined the effects of a repetitive collision in a season of NCAA Football Bowl Subdivision athletes (N=18) using multi-MRI modality, including diffusion tensor imaging (DTI), arterial spin labeling (ASL), resting-state functional MRI (rs-fMRI), susceptibility-weighted imaging (SWI) and T1-weighted structural imaging. No significant findings were found for DTI and cortical volumes. However, rsfMRI revealed significant changes in functional connections, including ASL results in global cerebral blood flow.

• Sollmann et al.(2018)[47] The authors investigated sex differences in microstructural changes of the brain measured by diffusion tensor imaging following exposure to repetitive HAE. This is done in a cohort of ice hockey players as part of the Hockey Concussion Education Project. Findings revealed significant differences between sexes in the right hemisphere’s SLF, IC, and CR. In these regions, FA decreased, while AD, RD, and MD increased for female subjects. Males did not show significant changes. Hence, this study suggests sex differences in structural alteration following exposure to repetitive HAE.

• Strauss et al.(2021)[88] The authors characterised the effects of soccer heading on WM microstructure and cognitive function through a comparison of amateur soccer players (N=246), non-contact/non-collision sports athletes (N=72), and healthy non-athletes (N=110). Furthermore, amateur soccer players are divided into quarters according to their exposure to heading. Athletes with no or low exposure to repetitive HAE exhibit a more significant decrease in RD and increase in FA and better performance on tasks of attention, processing speed, verbal memory, and working memory. Soccer players with high exposure did not differ from healthy non-athletes. Results were consistent with the notion of the beneficial effects of athletic conditioning on brain structure, which is attenuated by exposure to repetitive HAE.

• Tayebi et al.(2024)[56] The authors presented a novel method of integrating diffusion tensor metrics along the whole volume of the fibre bundle using a 3D mesh-morphing technique coupled with PCA for delineating rugby players (N=20) with non-contact athletes (N=12). Analyzing the whole volume of the WM tract provided clear delineation between rugby players and controls, which was not possible with traditional averaging methods.

• Tayebi et al.(2024)[55] The authors conducted a longitudinal study of collision sport athletes (N=36) to characterise season changes in DTI metrics as well as in comparison with controls (N=20) and subjects diagnosed with mTBI. No longitudinal changes were observed within collision sport athletes. However, a substantial difference in MD and AD was observed between collision sport athletes and controls. Pre- to post-season DTI changes within collision sport athletes also revealed a correlation with symptom scores.

• Wright et al.(2021)[65] The authors performed a longitudinal diffusion-weighted MRI study on male and female athletes playing Australian football. They sought to determine if microstructural changes in the brain detected by DTI are sex-dependent, considering different symptomology between males and females. The study reported more symptoms in male athletes with SRC than in female athletes. Thus, adding to the notion of considering sex better to understand the role of sex in SRC outcomes.

• Wu et al.(2020)[118] The authors studied the longitudinal trajectory of recovery using diffusion tensor imaging after sports-related mTBI of collegiate athletes who sustained SRC (N=82) compared to contact sport athletes (N=68) and non-contact sport athletes (N=69) scanned at the same time points. The study found significantly higher MD in concussed athletes compared with controls, which persisted even beyond asymptomatic. No significant difference was found between contact-sport and non-contact sport athletes.

• Yuan et al.(2018)[87] The authors investigated the longitudinal WM changes throughout two consecutive high school football seasons and explored the effect of jugular vein compression on WM alterations. Despite comparable exposure, non-collar athletes had reduced MD, AD, and RD in the first season, which did not exist among collar athletes. This reversed during the off-season. In the second season, the same trend was found still in the non-collar athletes but was localised in a spatially different region from the first season.

• Zimmerman et al.(2021)[101] The authors, in a longitudinal observational study, investigated the relationship between rugby participation and sub-acute head injuries among rugby players (N=44) along non-sporting controls (N=32), non-collision sport athletic controls (N=15), and longitudinally assessed controls (N=16). The non-acutely injured rugby players manifested abnormalities in FA and other diffusion indices, absent in non-collision sport athletes. Findings suggest an association of participation in elite adult rugby sports participation with changes in brain structure.

