## Supplementary Material for "How diffusion tensor imaging findings in subconcussive head impacts correlate with head kinematic exposures: a systematic review and quantitative synthesis"

### SUPPLEMENTARY MATERIALS

#### A. Optimised NIH quality rating tool and instructions (Lees, 2021)

**Modified Study Quality Assessment Tool from Lees, 2021.**

| Criteria | Maximum Scoring Allocation | Scoring guide |
|---|---|---|
| **1. Was the research question or objective in this paper clearly stated?** | 1 | 1 point was allocated if the article had clearly stated its aim or objective. |
| **2. Was the study population clearly specified and defined?** | 2 | 1 point was allocated if all demographic information was clearly reported (age and sex as minimum). No points were deducted if minor details were missing (*e.g.* years of exposure to sport, handedness, if educational level was indicated but exact years of education not specified). 1 point was deducted if demographic details were completely missing, or poorly reported.<br><br>1 point allocated if previous history of concussion or no history of concussion among subjects was reported as part of the demographic data. |
| **3. Were the cases clearly defined and differentiated from controls?** | 1 | For cross-sectional studies or longitudinal studies with cross-sectional arm: 1 point was allocated if a control group was included and clearly defined including demographics.<br><br>For longitudinal studies with no cross-sectional comparison: N/A |
| **4. Were all the subjects selected or recruited from the same or similar populations (including the same time period)? Were inclusion and exclusion criteria for being in the study pre-specified and applied uniformly to all participants?** | 2 | 2 points were allocated if the article had thorough discussion on recruitment and exclusion/inclusion criteria for exposure group and control group (if applicable).<br><br>1 point allocated if exclusion/inclusion were reported but they were not exhaustive. (*e.g.* no clear delineation between exposure and control group, no discussion of recruitment process, no statement of how consent was gathered)<br><br>0 if inclusion and exclusion criteria was not stated. |
| **5. Was a sample size justification, power description, or variance and effect estimates provided?** | 1 | 1 point was allocated if the study reported sample size in sufficient manner and provided reasons for exclusions of initial subjects in the final analysis.<br><br>0 if study did not specify how many subjects were included in the analysis. |
| **6. For the analyses in this paper, were the exposure(s) of interest measured prior to the outcome(s) being measured?** | 1 | For studies aiming to measure subject-specific or within group effect of sub-concussive injury or concussion to DTI measures: 1 point is allocated if DTI assessment was conducted at 2 or more time points.<br><br>For studies aiming to determine the effect of sub-concussive injury or concussion to DTI measures of exposed group in comparison to control group: Classed as N/A |
| **7. For exposures that can vary in amount or level, did the study examine different levels of the exposure as related to the outcome (e.g., categories of exposure, or exposure measured as continuous variable)?** | 2 | 1 point is allocated if valid measure of concussive symptomatology and/or measures of cognition were used including but not limited to ImPACT and SCAT or/and standardized tools to measures head impact were used (if applicable). No point if non-standardised measures were used or if results from any assessment of cognition, concussive symptomatology, or head impact exposure were not clearly reported.<br><br>1 point was allocated if study investigated the result of these measures in relation to DTI results. |
| **8. Were the exposure measures (independent variables) clearly defined, valid, reliable, and implemented consistently across all study participants?** | 2 | 2 points were allocated if within-study period concussion incidences (or its absence) were stated. For cases with within-study concussion, justification is stated on how it was dealt whether included or excluded in the analysis.<br><br>1 point only was allocated if information regarding within-study concussion, or its absence was stated but did not state if it was included or excluded in the analysis.<br><br>0 point was allocated if there was no information reported regarding a concussion diagnosis. |
| **9. Was the exposure(s) assessed more than once over time?** | 1 | For longitudinal studies with or without cross-sectional arm:<br>1 point was allocated if DTI was assessed longitudinally over time.<br><br>Classed as 'N/A' if DTI measurement was cross-sectional only. |
| **10. Were the outcome measures (dependent variables) clearly defined, valid, reliable, and implemented consistently across all study participants?[1]** | 3 | (See footnote for specific scoring criteria)<br><br>1 point was allocated if DTI results were reported accurately including direction (whether increase or decrease) and reference for stating such (whether before and after scan or with a reference group) |

| | | |
|---|---|---|
| | | 1 point was allocated if the DTI protocol was clearly defined and assessed what it aimed at measuring.<br><br>1 point was allocated if non-significant findings for DTI metrics studied are reported (if applicable). |
| **11. Was loss to follow-up after baseline 20% or less?** | 1 | For longitudinal studies, 1 point was allocated when loss to follow-up after baseline was 20% or less for the group under longitudinal arm of study. (*e.g.* exposed group, control group, or both)<br><br>Classified as 'N/A' if DTI assessment was cross-sectional. |
| **12. Were key potential confounding variables measured and adjusted statistically for their impact on the relationship between exposure(s) and outcome(s)?** | 1 | 1 point was allocated if minimum of one key confounding variable that could have impacted the DTI results was controlled.<br><br>0 points allocated if no key confounding variables were controlled. |

Q10 assessed the DTI protocols in detail relative to the aims of each study.

- Sufficiency of reported DTI parameters including MRI parameters (number of diffusion direction, b-values, head coil, type of sequence, Echo Time, Repeat Time, Field of View, Number of Slices, Gaps or No Gaps, Voxel Dimension, how B0 was or were acquired) and clear mention of preprocessing software, pipeline, and DTI metrics under study(FA, MD, AD, RD, etc). If not sufficiently reported, point is deducted.
- Deduction is made in case of 1) tractography used with non HARDI or other optimised protocol (e.g. isotropic resolution), 2) ROI or whole brain analysis including grey matter, 3) no mention of eddy and motion correction, 4) no description of registration process to anatomical or standard atlas image, 5) less than 30 gradient directions, or 6) poor rational of selection of ROIs or whole brain analysis.
- Deduction is made in case study: 1) Did not include outcomes that correctly measure the stated aims (e.g., aim was measuring FA in a set of selected ROI and other outcomes are added without rationale), 2) No corrections for multiple comparisons or analytic pipelines that do not reduce the total number of comparisons (e.g., TBSS, or selected ROIs), or 3) Protocol not consistently applied to all participants.

*Note.* DTI = diffusion tensor imaging; ImPACT = Immediate Post-Concussion Assessment and Cognitive Testing; N/A = not applicable; SCAT = The Sport Concussion Assessment Tool; SRC = sports related concussion; FA – Fractional Anisotropy; MD – Mean Diffusivity; AD – Axial Diffusivity; RD – Radial Diffusivity

#### B-1. Summary of methods of studies (M-Male; F-Female)

| NO | AUTHOR | NUMBER OF EXPERIMENTAL SUBJECTS AND DESCRIPTION | SPORT OF EXPERIMENTAL SUBJECTS | NUMBER OF CONTROL SUBJECTS AND DESCRIPTION | SPORT OF ATHLETIC CONTROL SUBJECTS | AGE (Years Old) | SEX (M/F) | HEAD IMPACT TOOL EMPLOYED | EXPOSURE METRICS |
|---|---|---|---|---|---|---|---|---|---|
| **1** | Asselin et al (2020) | 28 athletes | Football | 1 non-athlete subject (for parameter optimisation) | N/A | Mean: 19.8 | Experiment: 28/0 | HITS (Simbex) | 1. Device HIM<br>2. HIC, RIC, HITsp Time-weighted |
| **2** | Bahrami et al (2016) | 25 players | Football | N/A | N/A | Exp Range: 8-13<br>Exp Mean: 11.72 | Experiment: 25/0 | HITS | 1. Device HIM<br>2. Risk Weighted Exposure |
| **3** | Barber Fross et al (2019) | 21 High School Players | Football | 12 Youth Players | Football | Exp Mean: 17.33<br><br>Cont Mean: 13.08 | Experiment: 21/0<br>Control: 12/0 | GForce Tracker | 1. Impact Count<br>2. Device HIM |
| **4** | Bazarian et al (2012) | 9 athletes | Football<br>Ice Hockey | 6 controls | Mixture of orthopedic injury and non-injured | Exp Range: 16-18<br>Cont Range: 16-35 | Experiment: 9/0<br>Control: 5/1 | Self Record | 1. Impact Count |
| **5** | Bazarian et al (2014) | 10 players | Football | 5 players | Non-athlete | Exp Mean: 20.4<br>Cont Mean: 20.6 | Experiment: 10/0<br>Control: 5/0 | HITS | 1. Device HIM (LA, RA, GSI, HIC, HITsp)<br>2. Impact Count |
| **6** | Bazarian et al. (2024) | 30 players | Football | N/A | N/A | Mean: 19.5 | All male | HITS | 1. Impact Count Device HIM |
| **7** | Brett et al. (2021) | 75 players | Football | 46 non-contact sport athletes | Not provided | Mean: 18.52 | Mixed | Years of Participation | N/A |
| **8** | Champagne et al (2019) | 33 players | Football | 20 players | Football | Longitudinal Mean: 20.3<br>Cross-Sectional Mean: 19.6 | Longitudinal: 33/0<br>Cross-Sectional 20/0 | gForce Tracker Accelerometers | 1. Strain Metrics<br>2. Device HIM<br>3. Impact Count |
| **9** | Chen et al. (2023) | 36 players | Football, Soccer, Lacrosse | 45 non-contact sport athletes | Baseball, Cross Country, Track and Field, Basketball | Exp Mean: 19.60<br>Control Mean: 19.90 | Experiment: 36/0<br>Control: 45/0 | N/A | N/A |
| **10** | Chen et al (2022) | 24 players with SRC | Football | 26 players with Repetitive Head Impact<br><br>28 Non-contact-sport control athletes | Players with RHI (RHI): Football<br><br>Non-contact(HC): Baseball Field Event Cross-Country | Exp Mean: 19.7<br><br>Cont1 Mean: 19.3<br>Cont2 Mean: 19.7 | Experiment: 24/0<br>RHI: 26/0<br>HC: 28/0 | N/A | N/A |
| **11** | Chrisman et al (2016) | 17 players. | Soccer | N/A | N/A | Exp Mean: 12.6 | Experiment: 10/7 | xPatch | 1. Impact Count with Severity<br>2. Device HIM |

| | | | | | | | | | |
|---|---|---|---|---|---|---|---|---|---|
| **12** | Chun et al (2015) | 34 players | Football | 9 athletes | Non-collision athletes | Exp Range: 14-18<br><br>Cont Range: 14-18 | Experimental: 34/0<br>Control: 9/0 | HITS | 1. Impact Count<br>2. Device HIM |
| **13** | Churchill et al (2020) | 12 Concussed Athletes | Volleyball<br>Ice Hockey<br>Field Hockey<br>Football<br>Rugby<br>Basketball | 44 Athletic Controls | Volleyball<br>Ice Hockey<br>Field Hockey<br>Soccer<br>Football<br>Rugby<br>Basketball<br>Lacrosse | Exp Mean: 20.3<br><br>Cont Mean: 20.0 | Experiment: 20/24<br>Control: 7/5 | N/A | N/A |
| **14** | Davenport et al (2014) | 24 players | Football | N/A | N/A | Exp Mean: 16.9 | Experiment: 24/0 | HITS | 1. Device HIM<br>2. Risk Weighted Exposure |
| **15** | Davenport et al (2016) | 14 players. | Football | N/A | N/A | Exp Mean: 16.9 | Experiment: 24/0 | HITS | 1. Risk Weighted Exposure |
| **16** | DiCesare et al (2020) | 46 athletes (Split into Test and Train) | Soccer | N/A | N/A | Mean: 16.0 | Experiment: 0/46 | Head Mounted Sensors<br><br>Accelerometer (X2 Biosystems) | 1. Impact Count (Threshold Based, Heuristic, ML-Based) |
| **17** | Diekfuss et al. (2021) | 107 collar-group | Football | 106 non-collar group | Football | Mean: 15.8 (For all subjects prior to exclusion) | Collar: 107/0<br>Non-collar: 106/0 | CSx | 1. Impact Count<br>Device HIM |
| **18** | Diekfuss et al. (2021) | 117 players | Football | N/A | N/A | Mean: 16.41 | All male | CSx | 1. Impact Count<br>Device HIM |
| **19** | Dudley et al (2020) | 128 players | Soccer | N/A | N/A | N/A | Experiment: 0/128 | Accelerometer (CSx Systems) | 1. Impact Count with Severity |
| **20** | Gajawelli et al (2013) | 11 Athletes | Contact Sports | 13 Athletes | Non-contact Sports | Exp Mean: 20.4<br><br>Cont Mean: 19.5 | Experiment: 11/0<br><br>Control: 13/0 | N/A | N/A |
| **21** | Gong et al (2018) | 16 players | Football | N/A | N/A | Range: 15-17 | Experiment: 16/0 | HITS (Riddell) | 1. Impact Count |
| **22** | Goubran et al(2023) | 49 athletes | Football | 24 players | Volleyball | Exp Mean: 19.11<br>Cont Mean: 19.57 | Experiment: 49/0<br><br>Control: 24/0 | High Position Based Impact Risk | Impact Count |
| **23** | Holcomb et al (2021) | 102 players | Football | 16 Non-collision sports control athletes | Baseball<br>Basketball<br>Swimming<br>Tennis<br>Soccer | Exp Mean: 11.71<br><br>Cont Mean: 11.00 | Experiment: 102/0<br><br>HC: 16/0 | HITS (Simbex) | 1. Strain Metrics |
| **24** | Hunter et al (2019) | 312 players | Soccer | 110 Non-athletic | N/A | Exp Mean: 25.9 | Experiment: 225/87 | HeadCount | 1. Impact Count (Heading) |
| **25** | Jang et al (2019) | 61 high school athletes | Football | 15 high school athletes | Track<br>Swimming<br>Cross-country<br>Basketball | Exp Mean: 16.6<br><br>Cont Mean: 16.5 | Experiment: 162/0 | HITS or xPatch | 1. Impact Count with Threshold |

| | | | | | | | | | |
|---|---|---|---|---|---|---|---|---|---|
| | | | | | | | Control: 19/0 | | |
| **26** | Kelley et al. (2021) | 47 players | Football | 16 non-contact sport athletes | Swimming, Track, Tennis | Mean: 12.1 (For all subjects prior to exclusion) | Control: 16/0 | HITS | 1. Impact Count<br>2. Device HIM RWE |
| **27** | Koerte et al (2012) | 17 players | Ice Hockey | N/A | N/A | Exp Range: 20-26<br>Exp Mean: 22.24 | Experiment: 17/0 | N/A | N/A |
| **28** | Kuzminski et al (2018) | 17 athletes | Football | N/A | N/A | Mean Age: 16 | Experiment: 17/0 | HITS (Simbex) | 1. Impact Count with Threshold |
| **29** | Lancaster et al. (2016) | 26 concussed athletes | Football | 26 non-injured athletes | Football | Exp Mean: 17.6<br><br>Control Mean: 18.0 | Experiment: 26/0 | N/A | N/A |
| **30** | Lancaster et al (2018) | 17 players with concussion | Football | 20 players without concussion | Football | Exp Mean: 17.5<br><br>Cont Mean: 17.9 | Experiment: 17/0<br><br>Control: 20/0 | N/A | N/A |
| **31** | Lao et al (2015) | 11 players | Contact Sport | N/A | N/A | N/P | Experiment: 11/0 | N/A | N/A |
| **32** | Manning et al. (2020) | 70 players | Rugby | 31 non-contact sport athletes | Rowing Swimming | Exp Mean: 19.95<br><br>Control Mean: 19.61 | Experiment: 0/70 | GFT3 | 1. Device HIM<br>2. HIC |
| **33** | Marchi et al (2013) | 37 Players | Football (College) | N/A | N/A | Range of Means: | NP | Head Hit Index | 1. Impact Count |
| **34** | Mayinger et al (2018) | 15 athletes | Football | 5 non-athletes | N/A | Exp Mean: 20.0<br><br>Cont Mean: 20.93 | Experiment: 15/0 | N/A | N/A |
| **35** | McAllister et al (2013) | 80 athletes | Football<br>Ice Hockey | 70 athletes | Track<br>Crew<br>Nordic Skiing<br>etc. | Exp Mean: 19.5<br><br>Cont Mean: 19.0 | Experiment: 64/16<br><br>Control: 56/23 | HITS | 1. Impact Count<br>2. Device HIM |
| **36** | Merchant-Borna et al (2016) | 10 players | Football | N/A | N/A | Exp Mean: 20.7 | Experiment: 10/0. | HITS | 1. Device HIM<br>2. HIC, RIC, HITsp<br>2. Time-weighted |
| **37** | Merz et al (2020) | 75 athletes | Football<br>Ice Hockey | 79 athletes | Track<br>Crew<br>Nordic Skiing<br>etc. | Exp Mean: 19.07<br>Cont Mean: 19.51 | Experiment: 60/15<br><br>Control: 56/23 | HIT System | 1. Impact Count<br>2. Device HIM |
| **38** | Miller et al (2022) | 95 athletes | Football | N/A | N/A | Exp Mean: 11.9 | Experiment: 95/0 | HITS (Simbex) | 1. Risk Weighted Exposure<br>2. Strain Metrics |
| **39** | Muftuler et al (2020) | 96 Concussed Athletes | Football | 82 Non-concussed Athletes | Football | Exp Mean: 18.06<br><br>Cont Mean: 18.37 | Experiment: 96/0<br><br>Control: 82/0 | N/A | N/A |
| **40** | Mund et al. (2024) | 14 players | Football | N/A | N/A | Mean: 19.9 | Experimental: 14/0 | Video-verified Headers | Impact Count |
| **41** | Murdaugh et al (2018) | 16 athletes with SRC | Football | 12 athletes without SRC | Football | Exp Mean: 15.99<br>Cont Mean: 15.77 | Experiment: 16/0 | N/A | N/A |

| | | | | | | | | | |
|---|---|---|---|---|---|---|---|---|---|
| | | | | | | | Control: 12/0 | | |
| **42** | Myer et al (2016) - A | 21 Collar Players | Football | 21 Non-Collar Players | Football | Mean: 17.04 | Experiment: 21/0Control 21/0 | GForce Tracker | 1. Impact Count<br>2. Device HIM |
| **43** | Myer et al (2016) - B | 7 Collar Players | Hockey | 8 Non-collar Players | Hockey | Mean: 16.3 | Experiment: 7/0<br><br>Control: 8/0 | GForce Tracker | 1. Impact Count<br>2. Device HIM |
| **44** | Myer et al (2019) | 46 players (Divided to collar and noncollar group) | Soccer | N/A | N/A | Exp Range: 14-18 | Experiment: 0/46 | XPatch | 1. Impact Count<br>2. Device HIM |
| **45** | Nilsson et al. (2019) | 35 contact-sport players | Football | 12 non-contact sport athletes | Swimming | Exp Mean: 10.11<br>Control: 10.17 | Experimental: 35/0<br>Control: 12/0 | Shockbox | 1. Impact Count<br>Device HIM |
| **46** | Puvvada et al (2021) | 95 players | Football | N/A | N/A | Exp Range of Means: 11.4 to 12.3 | NP | HITS (MxEncoder) | 1. Impact Count<br>2. Risk Weighted Exposure<br>3. Time Weighted |
| **47** | Saghafi et al (2018) | 60 players | Football | N/A | N/A | Range: 9-18 | NP | HITS | 1. Risk Weighted Exposure |
| **48** | Schranz et al (2018) | 64 Players | Rugby(University) | N/A | N/A | Mean: 21 | Experiment: 0/64 | N/A | N/A |
| **49** | Slobonuv et al (2017) | 18 athletes | Football | N/A | N/A | Exp Mean: 21.6 | Experiment: 18/0 | BodiTrak | 1. impact Count with Severity |
| **50** | Sollmann et al (2018) | 25 players | Ice Hockey | N/A | N/A | Exp Mean:<br>Male: 21.7<br>Female: 19.2 | Experiment: 14/11 | N/A | N/A |
| **51** | Strauss et al (2021) | 246 Amateur Soccer Players | Soccer | 72 Non-collision athletes<br><br>116 Non-athletic controls | Baseball<br>Swimming<br>Tennis<br>Running/Track<br>Gymnastics<br>Rowing/Crew<br>Cycling<br>Dancing<br>Figure Skating | Exp Mean: 25.48<br><br>Control 1 Mean: 22.76<br><br>Control 2 Mean: 28.96 | Experiment: 174/72<br><br>Control 1: 30/42<br><br>Control 2: 63/53 | HeadCount | 1.Impact Count |
| **52** | Tayebi et al (2024) | 20 Players | Rugby | 12 Players | Non-contact Sports | Range: 14-18 | Experiment: 20/0<br><br>Control: 12/0 | Bespoke | 1. Device HIM |
| **53** | Tayebi et al. (2024) | 10 Concussed athletes<br>35 Non-concussed athletes | Rugby | 20 non-contact sport athletes | Basketball, Track and Field, Rowing, Outrigger Canoe | Concused Mean:15.6<br>Non-concussed Mean:16.22<br>Control: 16.6 | All male | HitIQ | Device HIM |
| **54** | Wright et al (2021) | 14 concussed athletes | Football | 16 nonconcussed athletes | Football | Exp Men Mean: 22.1<br>Exp Women Mean: 24.2 | Experiment: 8/6 | N/A | N/A |

| | | | | | | | | | |
|---|---|---|---|---|---|---|---|---|---|
| | | | | | | Cont Men Mean: 24.7<br>Cont Women Mean: 22.1 | Control:<br>9/7 | | |
| **55** | Wu et al (2020) | 82 Concussed Athletes | Football<br>Soccer<br>Lacrosse Team | 68 Contact Sport Athletes<br><br>69 Non-contact Sport Athletes | Baseball<br>Softball<br>Basketball<br>Tract and Field<br>Cross Country | Exp Mean: 18.87<br><br>Cont 1 Mean: 18.82<br>Cont 2 Mean: 19.20 | Experiment:<br>69/13<br>Control 1:<br>54/14<br>Control 2:<br>55/14 | N/A | N/A |
| **56** | Yuan et al (2018) | 42 athletes | Football | N/A | N/A | Exp: 16-17.9<br><br>Cont: 15.5-17.8 | Experiment:<br>42/0 | GForce Tracker Accelerometers | 1. Impact Count with Severity<br>2. Device HIM |
| **57** | Zimmerman et al (2021) | 44 active players | Rugby | 15 non-collision sport athletes<br><br>32 non-sporting controls<br><br>16 longitudinally assessed controls | Swimming<br>Water Polo<br>Rower<br>Cyclist<br>Weight Lifter | Exp Range: 16-45<br><br>Control Range: 16-45 | Experiment:<br>41/3 | N/A | N/A |

#### B-2. Summary of methods of studies (NP – Not provided, B – B-value)

| AUTHOR | DTI METRIC | OTHER DIFFUSION METRICS | CLINICAL OR NEUROCOGNITIVE TESTS | SCANNER | COIL | SEQUENCE TYPE | B-VALUE (Directions) | TR/TE (ms) | FOV (mmxmm) | MATRIX | RESOLUTION | THICKNESS | SLICES | SOFTWARE USED FOR DTI |
|---|---|---|---|---|---|---|---|---|---|---|---|---|---|---|
| Asselin et al (2020) | FA | N/A | SCAT<br>ImPACT<br>BESS | 3T Trio Siemens | 32-channel head coil | Spin Echo EPI | B1200(69)<br>B0(10) | 9,100/89 | NP | NP | 2x2x2 mm | NP | NP | FSL<br>MountDoom |
| Bahrami et al (2016) | FA | N/A | N/A | 3T Skyra Siemens | 32-channel head and neck coil | 2D Single Shot EPI | B1000(15)<br>B2000(15)<br>B0(10) | 10500/99 | NP | NP | 2.2 x 2.2 mm | 3mm | 54 | AFQ |
| Barber Fross et al (2019) | FA<br>MD<br>AD<br>RD | N/A | N/A | 3T Achieva Philip | 32-channel head coil | Single-Shot Spin-Echo EPI | B1000(61)<br>B0(7) | 9000/83 | 256x256 | 128x128 | 2x2mm | 2mm | 72 | FSL |
| Bazarian et al (2012) | FA<br>MD | N/A | SAC<br>ImPACT | 3T Trio Siemens | NP | Single-Shot Spin Echo EPI | B700(60)<br>B0(1) | 8000/89 | NP | NP | 2x2x2 mm | NP | NP | FSL |
| Bazarian et al (2014) | FA<br>MD | N/A | ImPACT<br>BESS<br>WBB | 3T Trio Siemens | NP | Single-shot SE-EPI | NP | NP | NP | NP | NP | NP | NP | FSL<br>Custom C++ and Matlab |
| Bazarian et al. (2024) | FA<br>MD<br>AD<br>RD | N/A | SCAT | 3T Siemens Magnetom | 64-channel head coil | Single-shot Spin-echo EPI | B1200(60)<br>B0(3) | 8000/89 | NP | NP | 2x2x2 mm | 2mm | NP | TORTOISE<br>ANTS<br>FSL |
| Brett et al. (2021) | FA<br>MD<br>AD<br>RD | KFA<br>MK<br>RK<br>AK | N/A | 3T MR750 GE | 32-channel | Single Shot Spin-Echo EPI | B1000(30)<br>B2000(30)<br>B0 | NP | NP | NP | 3.0mm Isotropic voxels | NP | NP | FSL<br>TBSS<br>DKE Software |
| Champagne et al (2019) | FA | N/A | N/A | 3T Magnetom Tim Trio Siemens | 32-channel receiver head | NP | B1000(30)<br>B0(4) | 7800/95 | 256x256 | 128x128 | 2mm isotropic | 2mm | 60 | FSL |
| Chen et al (2022) | FA<br>MD<br>AD<br>RD | MK<br>AK<br>RK<br>AWF<br>Daxon<br>Extra Axonal Axial<br>Extra Axonal Radial | SCAT3 | 3T Magnetom Prisma Scanner | NP | NP | B1000,2000(30)<br>B0(8) | 7900/98 | 243 x 243 | 90x90 | 2.7mm Isotropic | NP | 64 | MRTrix<br>FSL<br>in-house Matlab R2019b,<br>TBSS |
| Chen et al. (2023) | FA<br>MD | MK<br>AWF<br>Daxon<br>De<br>De | N/A | 3T Scanner (Center-specific) | Not provided | NP | B1000, B2000,<br>B0(8) | 7900/98 | 243x243 | 90x90 | 2.7mm Isotropic Voxels | NP | NP | FSL<br>TBSS<br>DESIGNER |
| Chrisman et al (2016) | FA | N/A | Neuropsychological Battery including ImPACT and other tests<br><br>22-item Post Concussion Symptom Scale | 3T Achieva Philips | 32-channel head coil | NP | B1000(64)<br>B0(6) | 7500/68.3 | NP | 112x112 | NP | 2mm | NP | TORTOISE |

| | | | | | | | | | | | | | | |
|---|---|---|---|---|---|---|---|---|---|---|---|---|---|---|
| Chun et al (2015) | FA | N/A | N/A | 3T Signa HDx GE | 16-channel brain array | Spin Echo EPI | B1000(30) | 12000/83.6 | 240x240 | NP | 2.5 mm isotropic | NP | 46 | FSL |
| Churchill et al (2020) | FA<br>MD | N/A | SCAT | 3T Skyra Magnetom | Multi-channel head coil | NP | B700(30)<br>B0(9) | 7800/83 | 240x240 | 120x120 | 2.0x2.0 | 2mm | 66 | FSL |
| Davenport et al (2014) | FA<br>MD | CL<br>CP<br>CS | ImPACT | 3T Skyra Siemens | 20-channel head/neck coil | 2D Single-shot EPI | B1000(15)<br>B2000(15)<br>B0(10) | 10500/99 | NP | NP | 2.2x2.2 mm | 3mm | 54 | FSL<br>DTI-TK.<br>SPM8<br>JMP |
| Davenport et al (2016) | N/A | DKI Metrics | ImPACT | 3T Skyra Siemens | 32-channel human head/neck coil | 2D Single Shot EPI | B1000(15)<br>B2000(15)<br>B0(10) | 10500/99 | NP | NP | 2.2x2.2 mm | 3mm | 54 | FSL<br>DKE |
| DiCesare et al (2020) | FA<br>MD<br>AD<br>RD | N/A | N/A | 3T Achieva Philips | 32-channel head coil | NP | B1000(61)<br>B0(7) | 8788/97 | 256x256 | 128x128 | 2x2x2 mm | 2mm | 68 | NP |
| Diekfuss et al. (2021) | FA<br>MD<br>AD<br>RD | N/A | N/A | 3 scanners:<br>3T Philips Achieva<br>3T Philips Ingenia<br>3T Philips Ingenia Elition | 32-channel head coils | Spin-echo EPI | B1000(61)<br>B0(7) | 8600/97 | 256x256 | 128x128 | 2x2 mm | 2mm | 67 | FSL |
| Diekfuss et al. (2021) | FA<br>MD<br>AD<br>RD | N/A | N/A | 3 scanners:<br>3T Philips Achieva<br>3T Philips Ingenia<br>3T Philips Ingenia Elition | 32-channel head coils | Spin-echo EPI | B1000(61)<br>B0(7) | 8600/97 | 256x256 | 128x128 | 2x2 mm | 2mm | 67 | FSL |
| Dudley et al (2020) | N/A | Structural Connectivity | N/A | 3T Achieva, Ingenia, or Ingenia Elition Philips | 32-channel phased array head coil | Spin EPI | B1000(61)<br>B0(7) | 8600/97 | 256x256 | 128x128 | 2x2x2 mm | 2mm | 67 | FSL<br><br>Diffusion Toolkit |
| Gajawelli et al (2013) | FA<br>MD | N/A | N/A | 3T HDxT GE | NP | Single-Shot Spin-Echo EPI | B1000(25)<br>B0(5) | 15.3/87 | 256x256 | NP | 2x2x2 mm | NP | NP | FSL<br>Tensor Toolkit<br>MedINRIA |
| Gong et al (2018) | FA<br>MD | MK | N/A | 3T MR750 GE | 8-channel head coil | NP | B1000/2000(31)<br>B0(2) | 10000/96 | NP | 128x128 | 2x2x2 mm | NP | 71 | DSI Studio |
| Goubran et al(2023) | FA<br>MD<br>AD<br>RD | ODI<br>ICVF<br>AWF<br>MK<br>AK<br>RK | SCAT | 3T MR750 GE | NP/81 | 2D Pulsed Gradient Spin Echo | B2500(60)<br>B800(30) | NP | NP | NP | 1.875x1.875x2 mm | NP | 70 | FSL<br>MRTRIX<br>AFQ<br>DESIGNER |
| Holcomb et al (2021) | FA<br>MD | CL<br>CS<br>CP | SCAT | 3T Skyra Siemens | 32-channel head coil | NP | B1000(15)<br>B2000(15) | P1: 10500/99<br>P2: 12600/100 | NP | NP | P1: 2.2mm x 2.2mm x 3.0 mm<br><br>P2: 2mm3 | NP | NP | FSL |

| | | | | | | | | | | | | | | |
|---|---|---|---|---|---|---|---|---|---|---|---|---|---|---|
| | | | | | | | | | | | isotropic voxel resolution | | | |
| Hunter et al (2019) | FA<br>MD<br>AD<br>RD | N/A | | 3T Achieva Philips | 32-channel head coil | 2D Single Shot Spin EPI | B800(32) | 10000/65 | NP | 128x120 | 2mm isotropic resolution | NP | 70 | FSL |
| Jang et al (2019) | FA<br>MD | N/A | N/A | 3T Signa HDxt GE | 16-channel brain array | 2D Single Shot Spin EPI | B1000(30)<br>B0(1) | 12000/83.6 | 240x240 | | 2.5x2.5mm | 2.5 mm | 46 | FSL |
| Kelley et al. (2021) | FA<br>MD | CL<br>CS<br>CP | N/A | 3T Siemens Skyra MRI Scanner | 32-channel head and neck coil | 2D Single Shot EPI | B1000(15)<br>B2000(15)<br>B0(10) | 10500/99 | NP | NP | 2.2 x 2.2 mm | 3mm | 54 | FSL |
| Koerte et al (2012) | FA<br>AD<br>RD<br>MD | N/A | SCAT2<br>ImPACT | 3T Achieva Philips | 8-channel head coil | NP | B600(60)<br>B0 | 7015/60 | NP | 100x100 | 2.2x2.2x2.2 mm | NP | 70 | FSL<br>3D Slicer |
| Kuzminski et al (2018) | FA | N/A | Computer-based Neurocognitive Assessment using CNS Vital Signs | 3T MR750 GE | 8-channel head coil | EPI | B1000(31) | 10000/96 | 256x256 | 128x128 | 2x2x2 mm | 2 mm | 71 | FSL |
| Lancaster et al. (2016) | FA<br>MD<br>AD<br>RD | MK<br>AK<br>RK | WTAR<br>SCAT-3<br>SAC<br>BESS | 3T MR750 GE | 32-channel head coil | Single-shot EPI | B2000(30)<br>B1000(30)<br>B0(4) | NP | NP | NP | 3.0mm Isotropic voxels | NP | NP | FSL, TBSS |
| Lancaster et al (2018) | FA<br>MD<br>AD<br>RD | MK<br>Kax<br>Krad | WTAR<br>SCAT<br>SAC<br>BESS | 3T MR750 GE | 32-channel head coil | Single-Shot Spin Echo EPI | B1000(30)<br>B2000(30)<br>B(4) | 5250/66.8 | NP | NP | NP | NP | NP | In-house software<br>FSL<br>TBSS |
| Lao et al (2015) | FA<br>MD | N/A | N/A | 3T HDxT GE | NP | Single Shot EPI | B1000(25) | NP | NP | NP | NP | NP | NP | MedINRIA |
| Manning et al. (2020) | FA<br>MD<br>AD<br>RD | N/A | SCAT-3 | 3T Prisma SIemens | 32-channel head coil | Spin-echo | B1000(64)<br>B0 | 7200/79 | 200x200 | 98x98 | | 2 mm | 64 | FSL, TBSS |
| Marchi et al (2013) | FA<br>MD | N/A | ImPACT | 3T Tim Trio Magnetom | | | B1200(60)<br>B0(10) | 10000/89 | | | 2x2x2 mm | | | Home-based FSL |
| Mayinger et al (2018) | FA<br>AD<br>RD | Trace | SCAT<br>ImPACT | 3T Tim Trio Siemens | 32-channel matrix head coil | NP | B1200(60)<br>B0(10) | 10000/89 | NP | NP | 2x2x2 mm | NP | NP | FSL<br>3D Slicer |
| McAllister et al (2013) | FA<br>MD | N/A | CVLT-II<br>WRAT-4 | 3T Achieva Philips | 8-channel SENSE head coil | NP | B1000(46)<br>B0(1) | 8100-8600/76 | NP | NP | 2x2x2 mm | NP | NP | ExploreDTI<br>FreeSurfer<br>SPM |
| Merchant-Borna et al (2016) | FA | N/A | N/A | 3T Trio Siemens | NP | Single-shot SE-EPI | NP | NP | NP | NP | NP | NP | NP | FSL<br>Custom C++ and Matlab |
| Merz et al (2020) | FA<br>MD | N/A | D-KEFS<br>Gordon CPT<br>PASAT<br>WAIS-III<br>WRAT-4 | 3T Achieva Philip | 8-channel head coil | NP | B1000(46)<br>B0(1) | 8100-8600/76 | NP | NP | 2x2x2 mm | NP | NP | Freesurfer<br>TRACULA |
| Miller et al (2022) | FA<br>MD | CL<br>CP<br>CS | N/A | 3T Siemens Skyra MRI Scanner | 32-channel head/neck coils | 2D Single-Shot EPI | B1000(15),<br>B2000(15),<br>B0(10) | 10,500/99 | NP | NP | 2.2x2.2 mm | 3mm | 54 | FSL |

| | | | | | | | | | | | | | | |
|---|---|---|---|---|---|---|---|---|---|---|---|---|---|---|
| Muftuler et al (2020) | FA<br>MD<br>AD<br>RD | KFA<br>MK<br>KRAD<br>KAX | SCAT<br>SAC<br>BESS | 3T MR750 GE | 32-channel head coil | Single Shot Spin-Echo EPI | B1000(30)<br>B2000(30)<br>B0 | NP | NP | NP | 3mm isotropic | NP | NP | FSL |
| Mund et al. (2024) | FA<br>MD<br>AD<br>RD | N/A | TMT A/B<br>PASAT 1 and 2 | 3T Philips Ingenia | 32 channel head coil | NP | B1000(32)<br>B0(1) | 5340/84 | NP | 128x128 | 1.75x1.75 | 2.5mm | NP | FSL<br>TBSS |
| Murdaugh et al (2018) | N/A | Connectivity Metrics | ImPACT | 3T Trio Siemens | NP | Spin Echo EPI | B1000(30)<br>B0(5) | 9200/95 | NP | NP | 2mm isotropic resolution | NP | NP | FSL<br>DSI Studio |
| Myer et al (2016) - A | FA<br>MD<br>AD<br>RD | N/A | N/A | 3T Achieva Philips | 32-channel head coil | Spin EPI | B1000(61)<br>B0(7) | 9000/83 | 256x256 | 128x128 | 2x2 mm | 2mm | 72 | FSL |
| Myer et al (2016) -B | FA<br>MD<br>AD<br>RD | N/A | N/A | 3T Achieva Philip | 32-channel head coil | Spin EPI | B1000(61)<br>B0(7) | 9000/83 | 256x256 | 128x128 | 2x2 mm | 2mm | 72 | FSL |
| Myer et al (2019) | FA<br>MD<br>RD<br>AD | N/A | N/A | 3T Achieva Philips | 32-channel head coil | Spin EPI | B1000(61)<br>B0(7) | 8788/97 | 256x256 | 128x128 | 2x2x2 mm | 2mm | 68 | FSL |
| Nilsson et al. (2019) | FA | N/A | N/A | 3T Siemens AG | 20-channel head coil | NP | B1000(64) | 10600-10800/91 | 128x128 | 256x96.9% | 2x2 mm | 2mm | 70-74 | Siemens Neuro 3D |
| Puvvada et al (2021) | FA<br>MD | CL<br>CP<br>CS | N/A | 3T Siemens Skyra MRI Scanner | 20-channel head/neck coil | 2D Single-Shot EPI | B1000(15),<br>B2000(15),<br>B0(10) | Protocol 1: 10,500/99<br>Protocol 2: 12,600/100 | NP | NP | Protocol 1: 2.2x2.2x3.0mm<br>Protocol 2: 2.0x2.0x2.0mm | 3mm | 54 | DTI-TK |
| Saghafi et al (2018) | FA | N/A | N/A | 3T Skyra Siemens | NP | NP | B1000(15)<br>B0(10) | NP | NP | NP | 2.2x2.2x2.2 mm | NP | NP | FSL<br>DTI-TK<br>VBM8 toolbox |
| Schranz et al (2018) | FA<br>MD<br>AD<br>RD | N/A | SCAT3<br>ImPACT<br><br>MRS | 3T Tim Trio Magnetom | 32-channel head coil | Spin-echo Echo-Planar DTI sequence | B1000(64)<br>B0 | 7200/79 | 200x200 | 98x98 | | 2mm | 64 | FSL |
| Slobonuv et al (2017) | FA<br>MD<br>AD<br>RD | N/A | N/A | 3T Prisma SIemens | 32-channel head coil | Spin Echo EPI | B1000(30)<br>B0(7) | 9800/94 | NP | 110x110 | 2x2x2 mm | 2mm | 72 | FSL |
| Sollmann et al (2018) | FA<br>MD<br>AD<br>RD | N/A | imPACT | 3T Achieva Philip | 8-channel head coil | NP | B700(60)<br>B0(2) | 7015/60 | NP | 100x100 | 2.2mm isotropic | NP | 70 | FSL<br>3D Slicer |
| Strauss et al (2021) | FA<br>MD<br>AD<br>RD | N/A | Cogstate<br>ISL<br>ISRL<br>GMCT<br>IDN<br>ONB<br>TWOB | 3T Philips Achieva TX | 32-channel head coil | 2D Single-Shot EPI | B800(32) | 10,000/65 | NP | 128x120 | 2mm3 isotropic | NP | 70 | FSL, ART |
| Tayebi et al. (2024) | FA<br>MD<br>AD<br>RD | N/A | SCAT | 3T GE SIGNA Premier | 48-channel head coil | Spin-echo | B3000(20)<br>B2000(15)<br>B1000(15)<br>B0(4) | 4500/70 | 260x260 | 128x128 | 2x2 mm | 2mm | | FSL<br>MRTRIX3 |

| | | | | | | | | | | | | | |
|---|---|---|---|---|---|---|---|---|---|---|---|---|---|
| Tayebi et al (2024) | FA<br>MD<br>AD<br>RD | N/A | N/A | 3T SIGNA Premier | 48-channel head coil | Multi-shel Spin Echo EPI | B3000(20)<br>B2000(15)<br>B1000(15)<br>B0(4) | 4500/70 | 260x260 | 128x128 | 2x2x2 mm | NP | 80 | FSL<br>MRTRIX<br>TractSeg |
| Wright et al (2021) | FA<br>RD<br>AD<br>MD | N/A | SCAT | 3T Prisma Siemens | NP | NP | B3000(64)<br>B0 | 3400/79 | 256x256 | 128x128 | 2mm isotropic resolution | NP | NP | FSL<br>MRTRIX |
| Wu et al (2020) | FA<br>MD<br>AD<br>RD | N/A | SCAT<br>BESS<br>BSI | 3T Tim Trio Magnetom | 12 channel or 32 receiver-only head coil | Single-shot EPI | B1000(30)<br>B0(8) | NP | NP | NP | NP | NP | NP | FSL |
| Yuan et al (2018) | FA<br>MD<br>AD<br>RD | N/A | N/A | 3T Achieva Philips | 32-channel head coil | Spin EPI | B1000(61)<br>B0(7) | 9000/83 | 256x256 | 128x128 | 2x2 mm | 2mm | 72 | FSL |
| Zimmerman et al (2021) | FA<br>MD | NDI<br>ODI<br>ISOVF | Battery of Neuropsychological Tests<br><br>SCAT5<br>TOPF<br>TMT<br>D-KEFS Stroop<br>HVLT<br>BVMT | 3T Siemens Verio | 32-channel head coil | NP | B700(30),<br>B2000(60),<br>B0(6) | 5,000/85 | 256x256 | NP | 2mm3 isotropic voxels | NP | 66 | FSL, TBSS<br><br>Freesurfer<br><br>DTI-TK |

C. Bar chart of all DTI parameters, comparing A) subclinical HAE-exposed subjects with normal controls (non-contact sport athletes or non-athletes)  B) subclinical HAE-exposed subjects with mTBI-diagnosed or concussion-diagnosed subjects. Data is presented in 3 categories: Significant difference (significant difference between groups), no significant difference, and did not state (findings were not reported for the particular DTI metric).

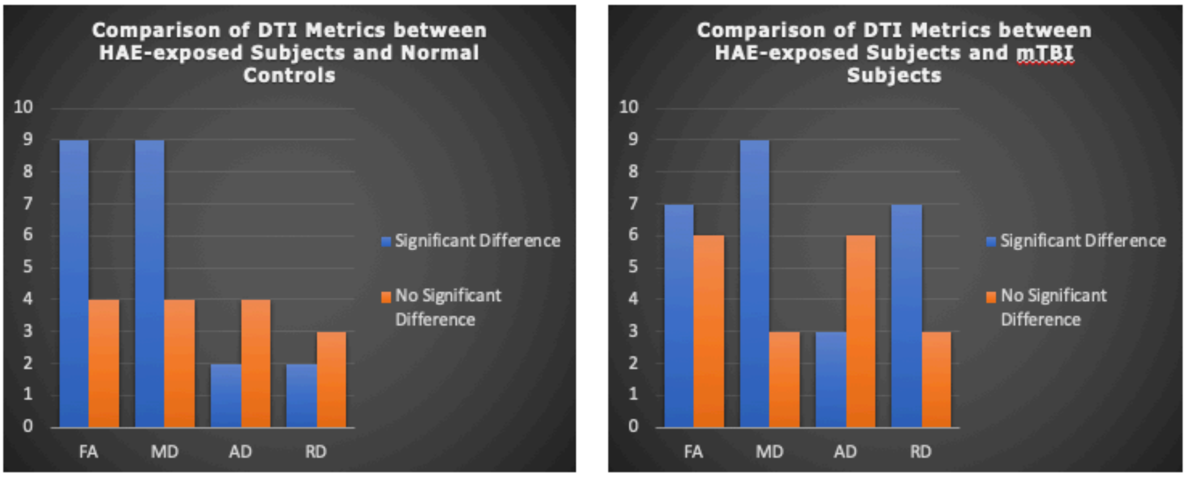

#### D. Study rating based on Appendix C: Modified Study Quality Assessment Tool from Lees, 2021.

| | Q1 | Q2 | Q3 | Q4 | Q5 | Q6 | Q7 | Q8 | Q9 | Q10 | Q11 | Q12 | Total Score | Maximum Possible. Score |
|---|---|---|---|---|---|---|---|---|---|---|---|---|---|---|
| Asselin et al (2020) | 1 | 2 | 1 | 2 | 1 | 1 | 2 | 2 | 1 | 3 | 1 | 1 | 18 | 18 |
| Bahrami et al (2016) | 1 | 2 | N/A | 2 | 1 | 1 | 2 | 1 | 1 | 3 | 1 | 1 | 16 | 17 |
| Barber Fross et al (2019) | 1 | 2 | N/A | 2 | 1 | 1 | 2 | 1 | 1 | 2 | 0 | 1 | 14 | 17 |
| Bazarian et al (2012) | 1 | 1 | 1 | 2 | 1 | 1 | 2 | 2 | 1 | 3 | 1 | 1 | 17 | 18 |
| Bazarian et al (2014) | 1 | 2 | 1 | 2 | 1 | 1 | 2 | 2 | 1 | 3 | 1 | 0 | 17 | 18 |
| Bazarian et al. (2024) | 1 | 2 | 1 | 2 | 1 | 1 | 2 | 2 | 1 | 3 | 1 | 1 | 18 | 18 |
| Brett et al. (2021) | 1 | 1 | 1 | 2 | 1 | 1 | 1 | 2 | 0 | 2 | 1 | 1 | 14 | 18 |
| Champagne et al (2019) | 1 | 2 | 1 | 2 | 1 | 1 | 2 | 2 | 1 | 3 | 1 | 0 | 17 | 18 |
| Chen et al (2022) | 1 | 2 | 1 | 1 | 1 | 1 | 1 | 2 | 1 | 3 | 1 | 1 | 16 | 18 |
| Chen et al. (2023) | 1 | 1 | 1 | 2 | 1 | 1 | 0 | 2 | 0 | 2 | 1 | 0 | 12 | 18 |
| Chrisman et al (2016) | 1 | 2 | 1 | 2 | 1 | N/A | 1 | 2 | N/A | 3 | 1 | 0 | 14 | 16 |
| Chun et al (2015) | 1 | 2 | N/A | 2 | 1 | 1 | 2 | 2 | 1 | 2 | 1 | 0 | 15 | 17 |
| Churchill et al (2020) | 1 | 1 | 1 | 1 | 1 | 1 | 2 | 2 | 1 | 3 | 1 | 0 | 15 | 18 |
| Davenport et al (2014) | 1 | 2 | 1 | 1 | 1 | 1 | 1 | 2 | 1 | 2 | 0 | 1 | 14 | 18 |
| Davenport et al (2016) | 1 | 2 | N/A | 2 | 1 | 1 | 2 | 2 | 1 | 2 | 1 | 1 | 16 | 17 |
| DiCesare et al (2020) | 1 | 2 | N/A | 2 | 1 | 1 | 2 | 2 | 1 | 2 | 1 | 1 | 16 | 17 |
| Diekfuss et al. (2021) | 1 | 2 | 1 | 2 | 1 | 1 | 2 | 2 | 1 | 3 | 1 | 1 | 18 | 18 |
| Diekfuss et al. (2021) | 1 | 2 | 1 | 2 | 1 | 1 | 1 | 2 | 1 | 3 | 1 | 1 | 17 | 18 |

| | | | | | | | | | | | | | |
|---|---|---|---|---|---|---|---|---|---|---|---|---|---|
| **Dudley et al (2020)** | 1 | 2 | N/A | 1 | 1 | 1 | 2 | 2 | 1 | 2 | 1 | 1 | 15 | 17 |
| **Gajawelli et al (2013)** | 1 | 1 | N/A | 1 | 1 | 1 | 2 | 2 | 1 | 3 | 0 | 1 | 14 | 17 |
| **Gong et al (2018)** | 1 | 1 | 1 | 2 | 1 | N/A | 0 | 1 | N/A | 2 | N/A | 0 | 9 | 15 |
| **Goubran et al(2023)** | 1 | 1 | N/A | 2 | 1 | 1 | 2 | 2 | 1 | 2 | 1 | 1 | 15 | 17 |
| **Holcomb et al (2021)** | 1 | 2 | 1 | 2 | 1 | 1 | 2 | 2 | 1 | 2 | 0 | 1 | 16 | 18 |
| **Hunter et al (2019)** | 1 | 2 | 1 | 2 | 1 | 1 | 2 | 2 | 1 | 3 | 1 | 1 | 18 | 18 |
| **Jang et al (2019)** | 1 | 2 | 0 | 2 | 1 | 1 | 1 | 1 | 1 | 3 | 1 | 1 | 15 | 18 |
| **Kelley et al. (2021)** | 1 | 1 | 1 | 2 | 1 | 1 | 2 | 2 | 1 | 2 | 0 | 1 | 15 | 18 |
| **Koerte et al (2012)** | 1 | 2 | 1 | 1 | 1 | 1 | 1 | 2 | 1 | 3 | 0 | 1 | 15 | 18 |
| **Kuzminski et al (2018)** | 1 | 2 | N/A | 2 | 1 | 1 | 2 | 1 | 1 | 3 | 0 | 0 | 14 | 17 |
| **Lancaster et al (2018)** | 1 | 2 | N/A | 2 | 1 | 1 | 2 | 2 | 1 | 3 | 1 | 1 | 17 | 17 |
| **Lancaster et al. (2016)** | 1 | 2 | 1 | 1 | 1 | 1 | 2 | 2 | 1 | 3 | 1 | 1 | 17 | 18 |
| **Lao et al (2015)** | 1 | 2 | 1 | 2 | 1 | 1 | 2 | 2 | 1 | 3 | 0 | 1 | 17 | 18 |
| **Marchi et al (2013)** | 1 | 1 | N/A | 1 | 1 | 1 | 0 | 1 | 1 | 1 | 0 | 0 | 8 | 17 |
| **Mayinger et al (2018)** | 1 | 2 | N/A | 1 | 1 | 1 | 2 | 1 | 1 | 1 | 1 | 1 | 13 | 17 |
| **McAllister et al (2013)** | 1 | 2 | 1 | 1 | 1 | 1 | 2 | 2 | 1 | 2 | 0 | 1 | 15 | 18 |
| **Merchant-Borna et al (2016)** | 1 | 2 | 1 | 2 | 1 | 1 | 2 | 2 | 1 | 3 | 1 | 1 | 18 | 18 |
| **Merz et al (2020)** | 1 | 2 | N/A | 1 | 1 | 1 | 2 | 2 | 1 | 3 | 1 | 0 | 15 | 17 |
| **Miller et al (2022)** | 1 | 2 | 1 | 2 | 1 | 1 | 2 | 2 | 1 | 3 | 1 | 1 | 18 | 18 |
| **Muftuler et al (2020)** | 1 | 2 | 1 | 2 | 1 | 1 | 2 | 1 | 1 | 1 | 1 | 1 | 15 | 18 |
| **Mund et al. (2024)** | 1 | 2 | N/A | 2 | 1 | 1 | 2 | 2 | 1 | 3 | 0 | 1 | 16 | 17 |

| | | | | | | | | | | | | | |
|---|---|---|---|---|---|---|---|---|---|---|---|---|---|
| **Murdaugh et al (2018)** | 1 | 2 | 1 | 2 | 1 | 1 | 2 | 2 | 1 | 3 | 1 | 1 | 18 | 18 |
| **Myer et al (2016) - A** | 1 | 2 | 1 | 2 | 1 | 1 | 2 | 2 | 1 | 2 | 1 | 1 | 17 | 18 |
| **Myer et al (2016) -B** | 1 | 2 | N/A | 2 | 1 | 1 | 2 | 1 | 1 | 3 | 0 | 1 | 15 | 17 |
| **Myer et al (2019)** | 1 | 2 | N/A | 2 | 1 | 1 | 2 | 1 | 1 | 3 | 0 | 1 | 15 | 17 |
| **Nilsson et al. (2019)** | 1 | 2 | 1 | 2 | 1 | 1 | 2 | 1 | 1 | 3 | 1 | 1 | 17 | 18 |
| **Puvvada et al (2021)** | 1 | 2 | N/A | 2 | 1 | 1 | 2 | 2 | 1 | 2 | 1 | 1 | 16 | 17 |
| **Saghafi et al (2018)** | 1 | 2 | 1 | 2 | 1 | 1 | 2 | 1 | 1 | 3 | 1 | 1 | 17 | 18 |
| **Schranz et al (2018)** | 1 | 2 | N/A | 1 | 1 | 1 | 2 | 1 | 1 | 1 | 1 | 1 | 13 | 17 |
| **Slobonuv et al (2017)** | 1 | 2 | 1 | 2 | 1 | 1 | 2 | 2 | 1 | 3 | 0 | 1 | 17 | 18 |
| **Sollmann et al (2018)** | 1 | 2 | N/A | 1 | 1 | 1 | 2 | 2 | 1 | 3 | 0 | 1 | 15 | 17 |
| **Strauss et al (2021)** | 1 | 1 | N/A | 2 | 1 | 1 | 2 | 2 | 1 | 3 | 0 | 1 | 15 | 17 |
| **Tayebi et al (2024)** | 1 | 2 | 1 | 2 | 1 | N/A | 2 | 2 | N/A | 3 | 0 | 1 | 15 | 16 |
| **Tayebi et al. (2024)** | 1 | 2 | 1 | 2 | 1 | 1 | 2 | 2 | 1 | 3 | 1 | 1 | 18 | 18 |
| **Wright et al (2021)** | 1 | 2 | 1 | 2 | 1 | 1 | 1 | 2 | 1 | 3 | 0 | 1 | 16 | 18 |
| **Wu et al (2020)** | 1 | 2 | 1 | 2 | 1 | N/A | 2 | 2 | N/A | 3 | 1 | 1 | 16 | 16 |
| **Yuan et al (2018)** | 1 | 2 | 1 | 1 | 1 | 1 | 2 | 2 | 1 | 2 | 1 | 1 | 16 | 18 |
| **Zimmerman et al (2021)** | 1 | 2 | N/A | 2 | 1 | 1 | 2 | 1 | 1 | 3 | 0 | 1 | 15 | 17 |
| **Manning et al. (2020)** | 1 | 2 | 1 | 2 | 1 | 1 | 2 | 2 | 1 | 3 | 0 | 1 | 17 | 18 |
